# Mental health of health care workers during the COVID-19 pandemic and evidence-based frameworks for mitigation: A rapid review

**DOI:** 10.1101/2021.01.03.21249166

**Authors:** Ajay Major, Fay J. Hlubocky

## Abstract

**Background:** The ongoing COVID-19 pandemic has profoundly affected the mental health of health care workers (HCWs), and optimal strategies to provide psychological support for HCWs are not currently established.

**Aims:** To rapidly review recently-published literature on the mental health of HCWs during the COVID-19 pandemic.

**Methods:** Query of all quantitative research through the PubMed database on the mental health of HCWs during the COVID-19 pandemic which utilized validated mental health instruments. 723 articles were screened and 87 articles were included.

**Results:** Nearly all included studies were cross-sectional, survey-based assessments of the prevalence of and risk factors for mental illness. Only one interventional study was identified. Prevalence of mental health outcomes varied widely: 7.0-97.3% anxiety, 10.6-62.1% depression, 2.2-93.8% stress, 3.8-56.6% post traumatic stress, 8.3-88.4% insomnia, and 21.8-46.3% burnout. Risk and protective factors were identified in personal and professional domains, including degree of COVID-19 exposure, adequacy of protective equipment, and perception of organizational support.

**Conclusions:** The myriad risk factors for poor mental health among HCWs suggests that a comprehensive psychosocial support model with individual- and organization-level interventions is necessary. Further longitudinal research on specific evidence-based interventions to mitigate adverse mental health outcomes among HCWs is urgently needed as the pandemic continues.

## Introduction

The COVID-19 pandemic is an unprecedented phenomenon that has profoundly affected health care delivery on a global scale. As the pandemic has evolved and various regions in the United States as well as across the world have experienced cycles of outbreaks and recovery, the laypress and academic literature have increasingly focused on the impact of the pandemic on the mental health of health care workers (HCWs).^1^ Research from previous epidemics, particularly the Severe Acute Respiratory Syndrome (SARS) epidemic of 2003, has demonstrated high rates of both acute and chronic mental health among HCWs who cared for patients with SARS.^2,3^ With headlines detailing the ‘moral injury’ of clinicians and the effects of repeated psychological trauma from caring for patients with COVID-19,^4^ it is critically important to review the current literature base to inform our approach towards caring for the psychological needs of HCWs, which may be different from previous infectious epidemics. In this manuscript, we perform a rapid review of literature on the mental health of HCWs during the ongoing COVID-19 pandemic, and evaluate evidence-based interventions to mitigate adverse mental health outcomes in HCWs.

## Materials and methods

In this rapid review of quantitative research studies assessing the mental health of HCWs during the COVID-19 pandemic, we searched for potential eligible articles on the PubMed online database on September 14, 2020 using the following search term: (COVID*) AND (health worker OR healthcare worker OR healthcare professional OR health professional) AND (mental OR psych*). Inclusion criteria were as follows: primary research on the mental health or psychological effects of the COVID-19 pandemic on HCWs, use of a validated psychometric instrument, published in a peer-reviewed journal, and written in English. Exclusion criteria included systematic reviews, meta-analyses, qualitative studies, pre-prints, and studies of only dentists. Studies which included both the general public and HCWs were included if the outcomes for HCWs could be easily distinguished from the general public. Interventional studies were only included if specific validated instrument outcomes resulting from the intervention were studied. Data extraction was performed in Microsoft Excel and relevant information, including country, dates of survey assessment, sample size, types of HCWs workers studied, instruments used, and outcomes were noted. Specific topic areas and themes of included studies were combined and presented as follows.

## Results

The database search identified 723 articles, as shown in the PRISMA diagram in **Figure 1**. After screening titles and abstracts, 581 were excluded based on the aforementioned inclusion and exclusion criteria. A total of 87 studies met inclusion criteria and were included for qualitative synthesis after full-text reading. Of these, 86 were epidemiological studies of HCWs and 1 was an interventional study. The full list of included articles are summarized in **Table 1**.

**Table 1.**
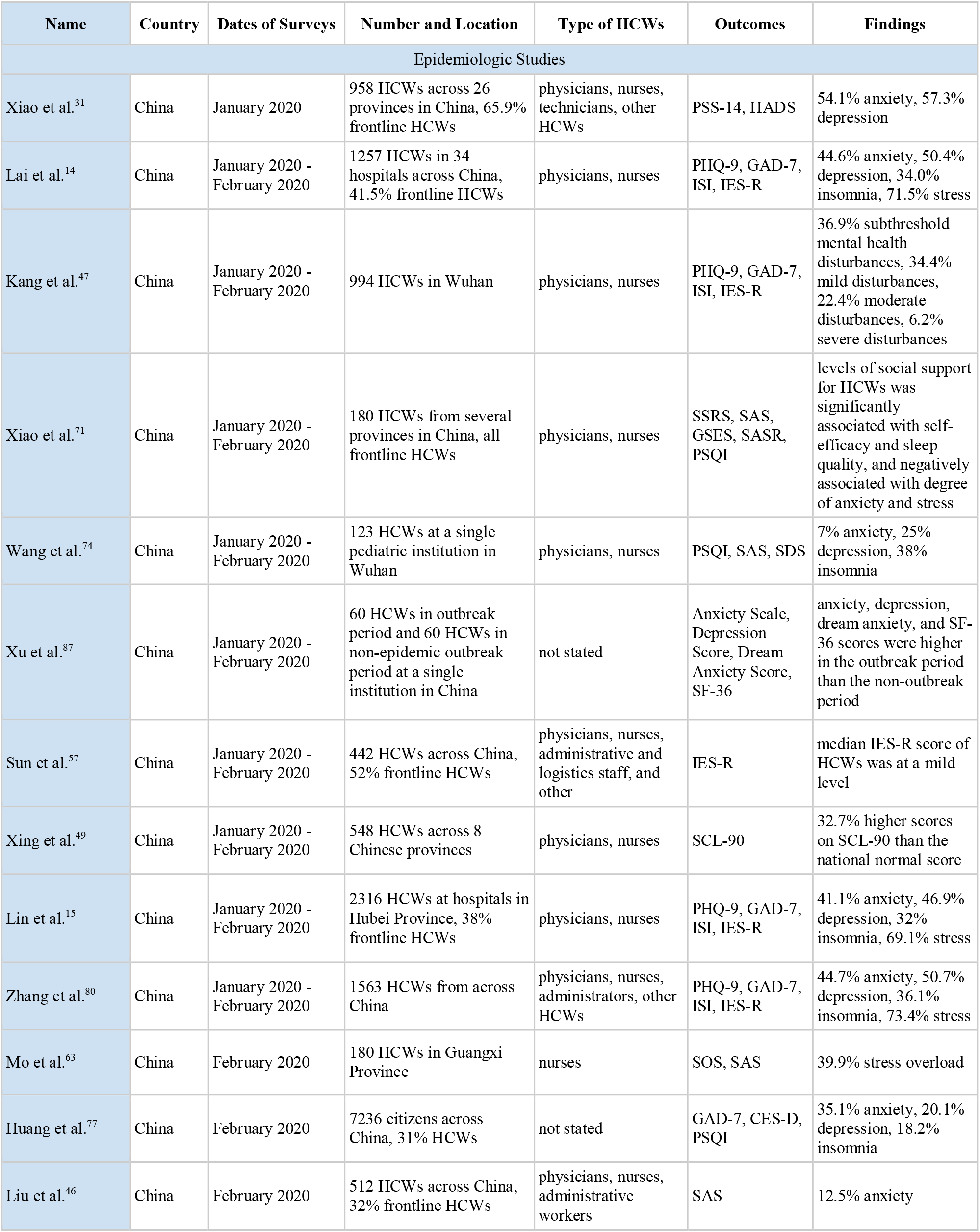
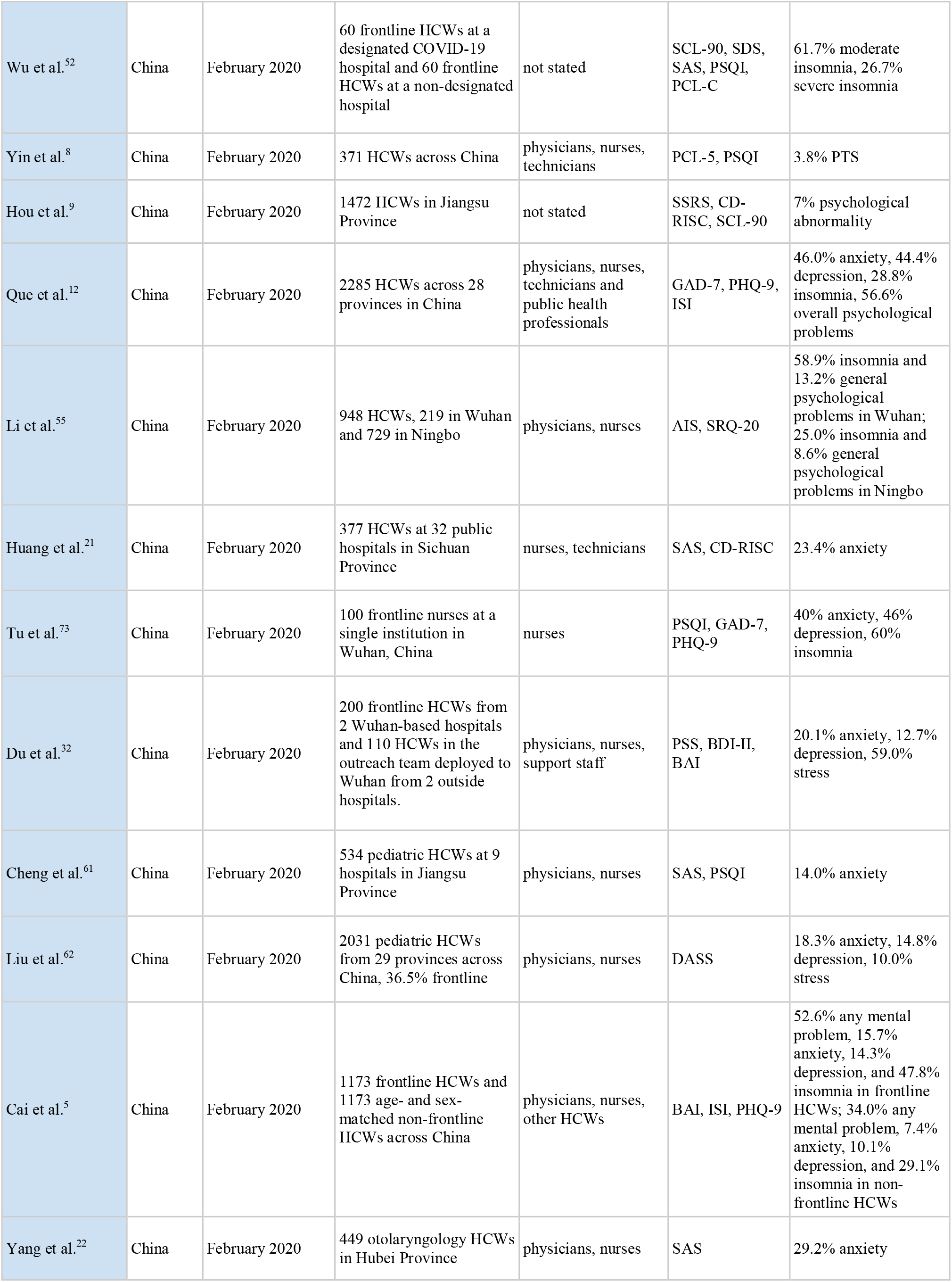
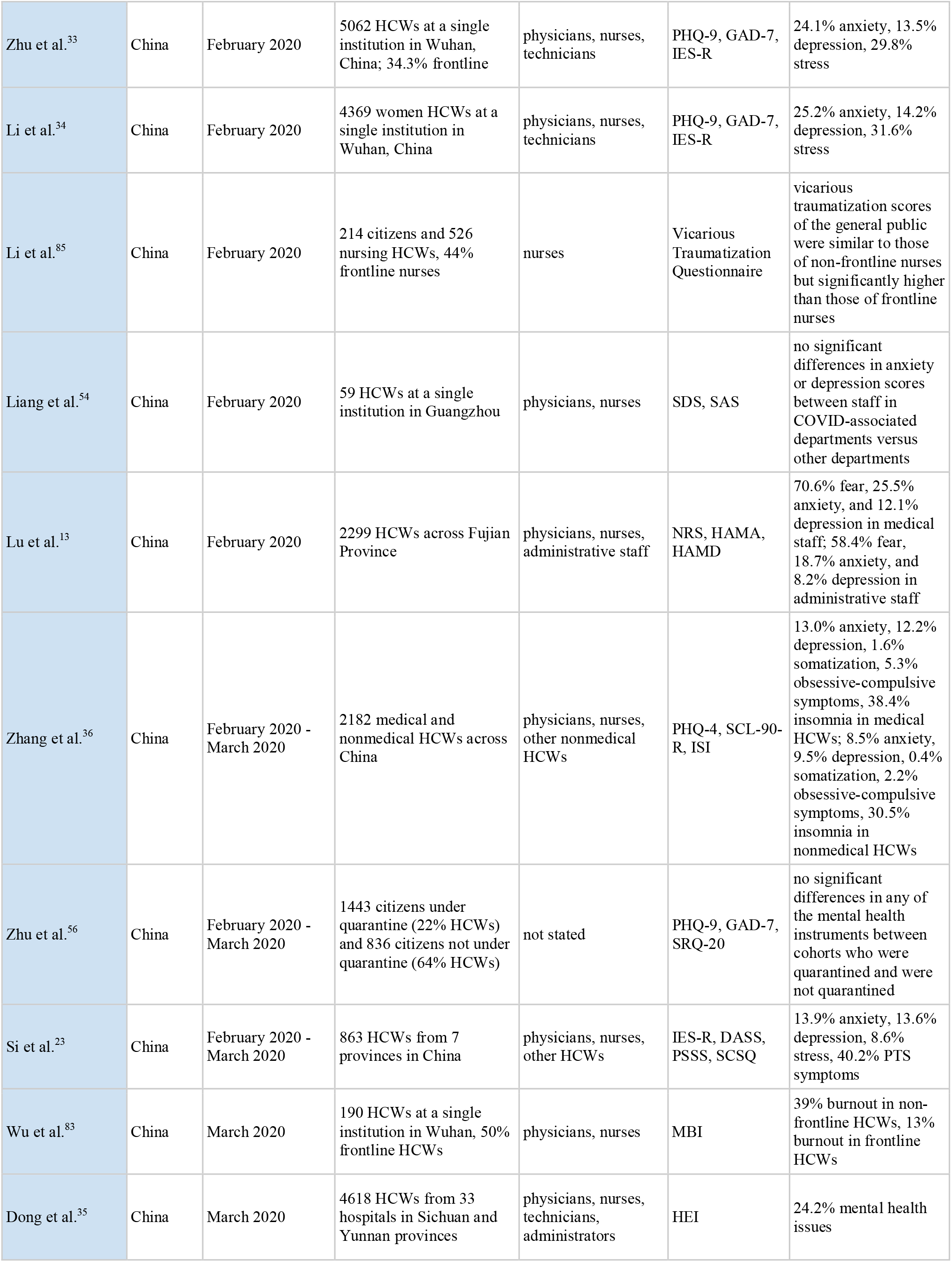
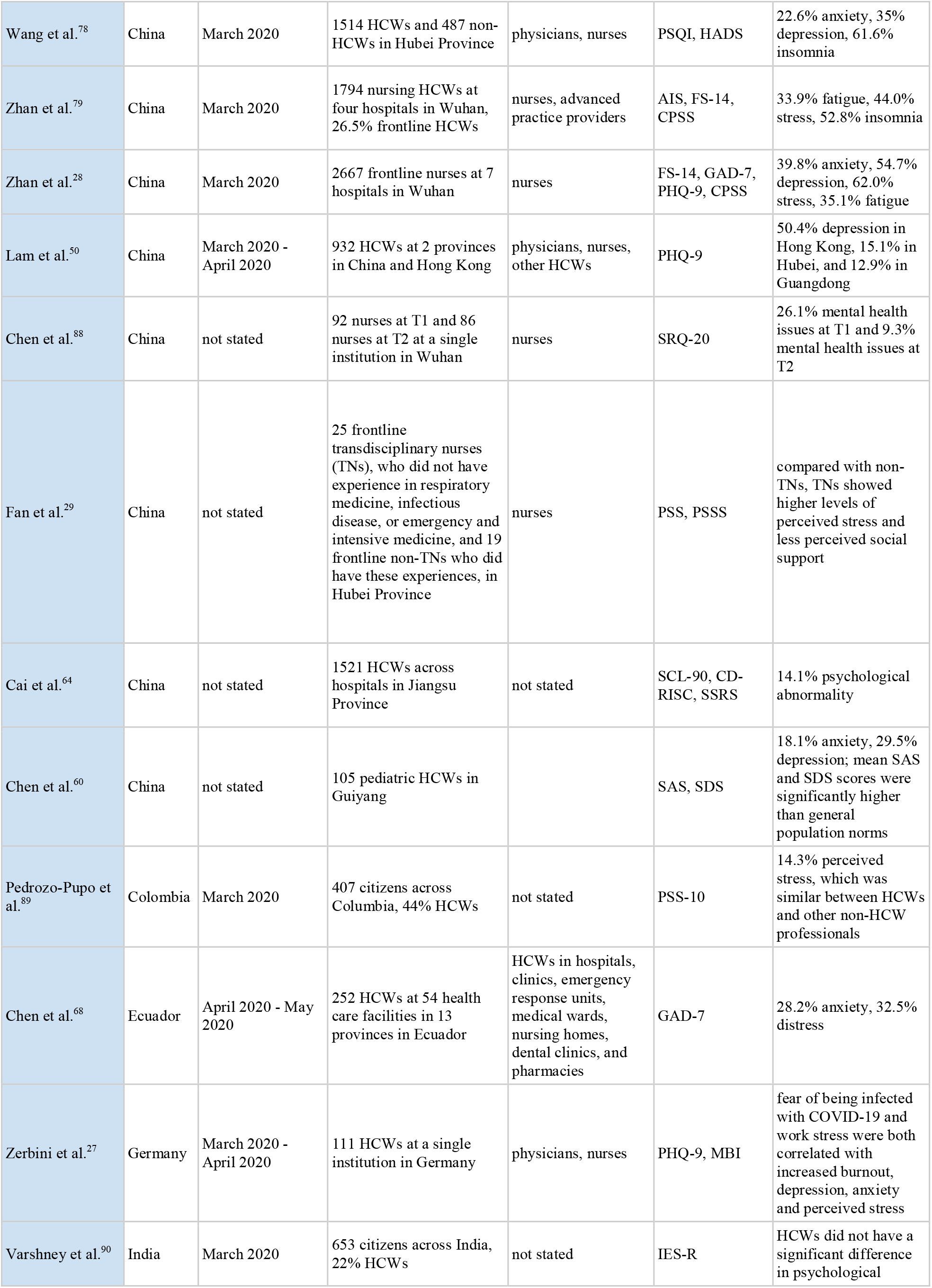
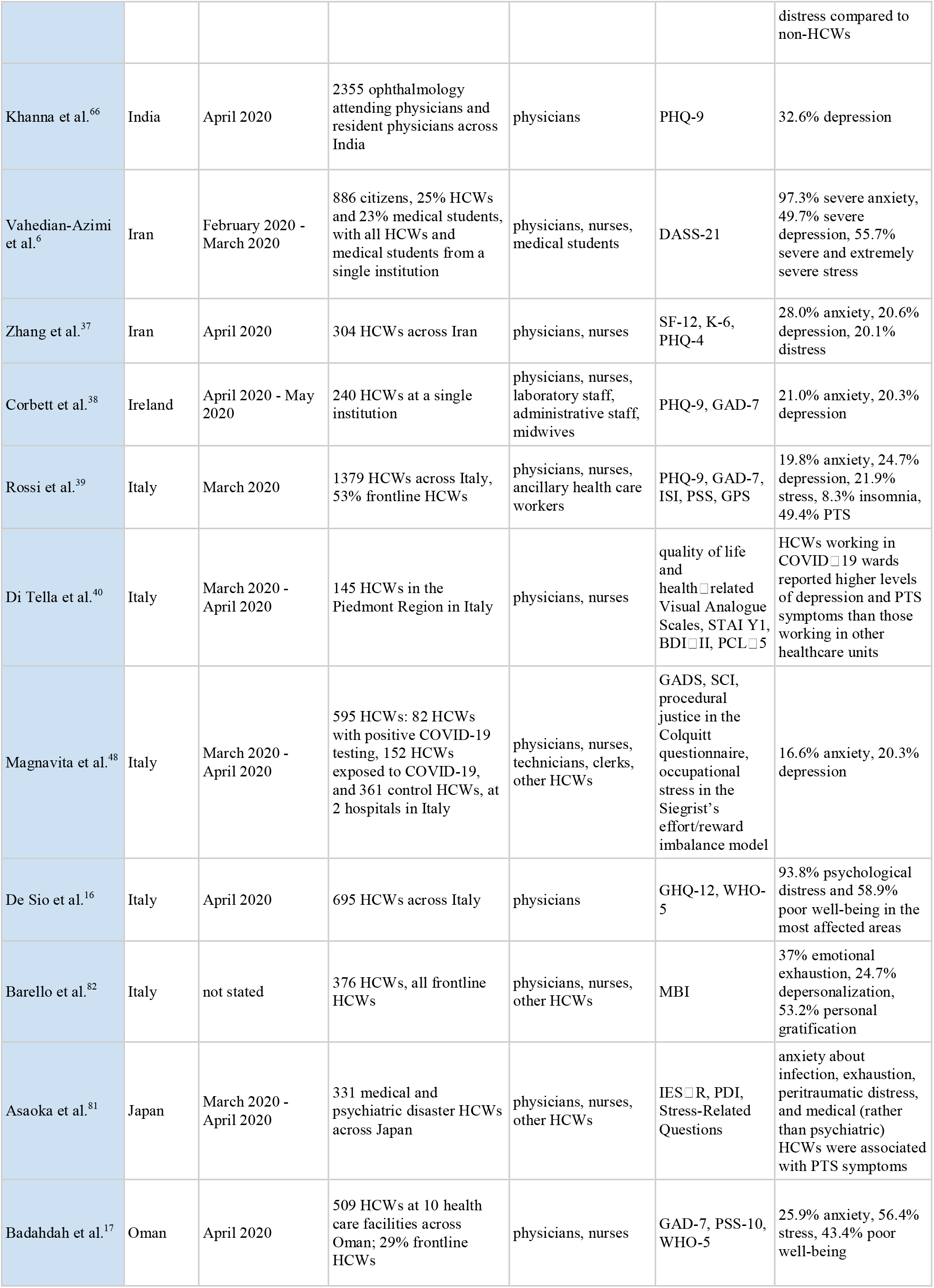
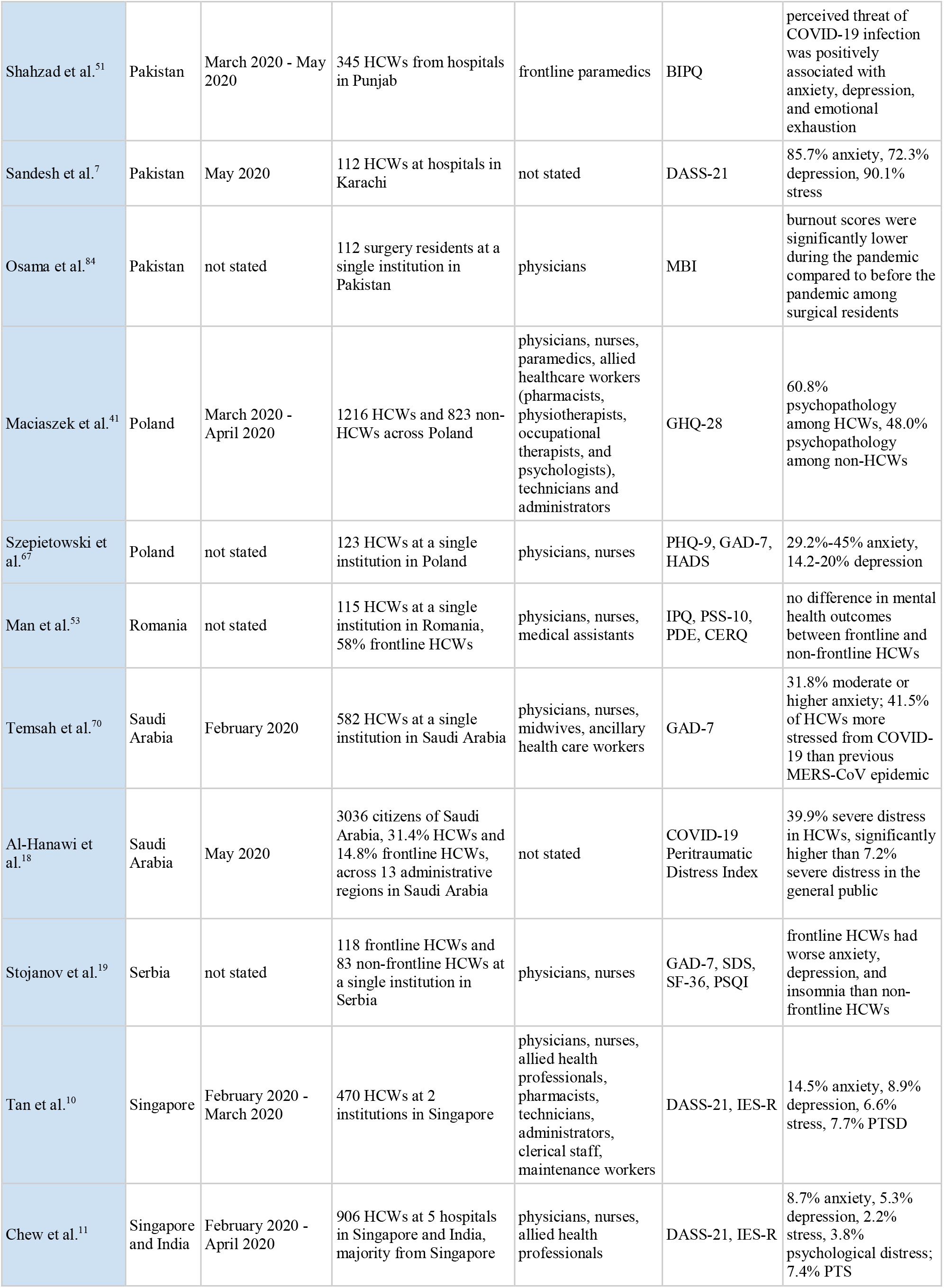
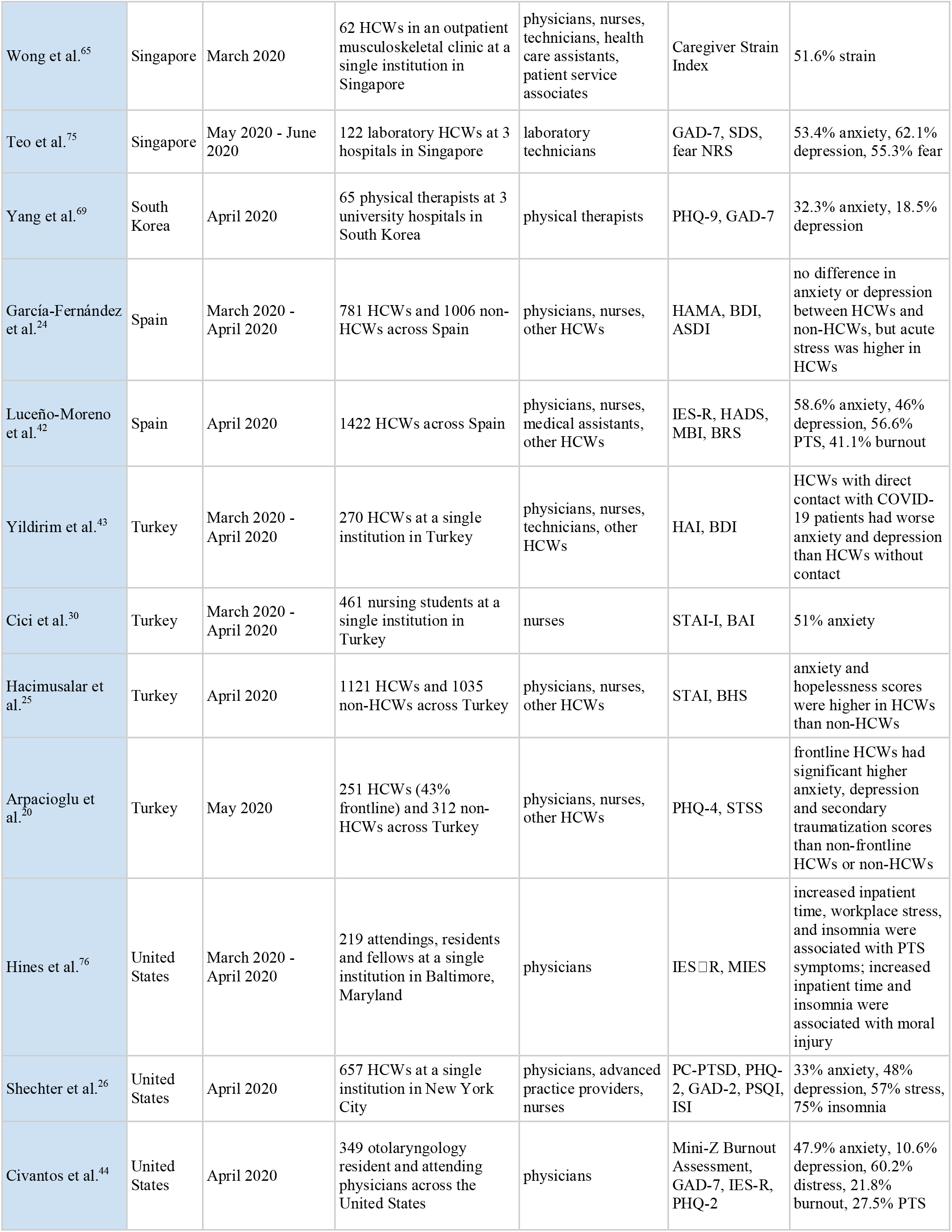
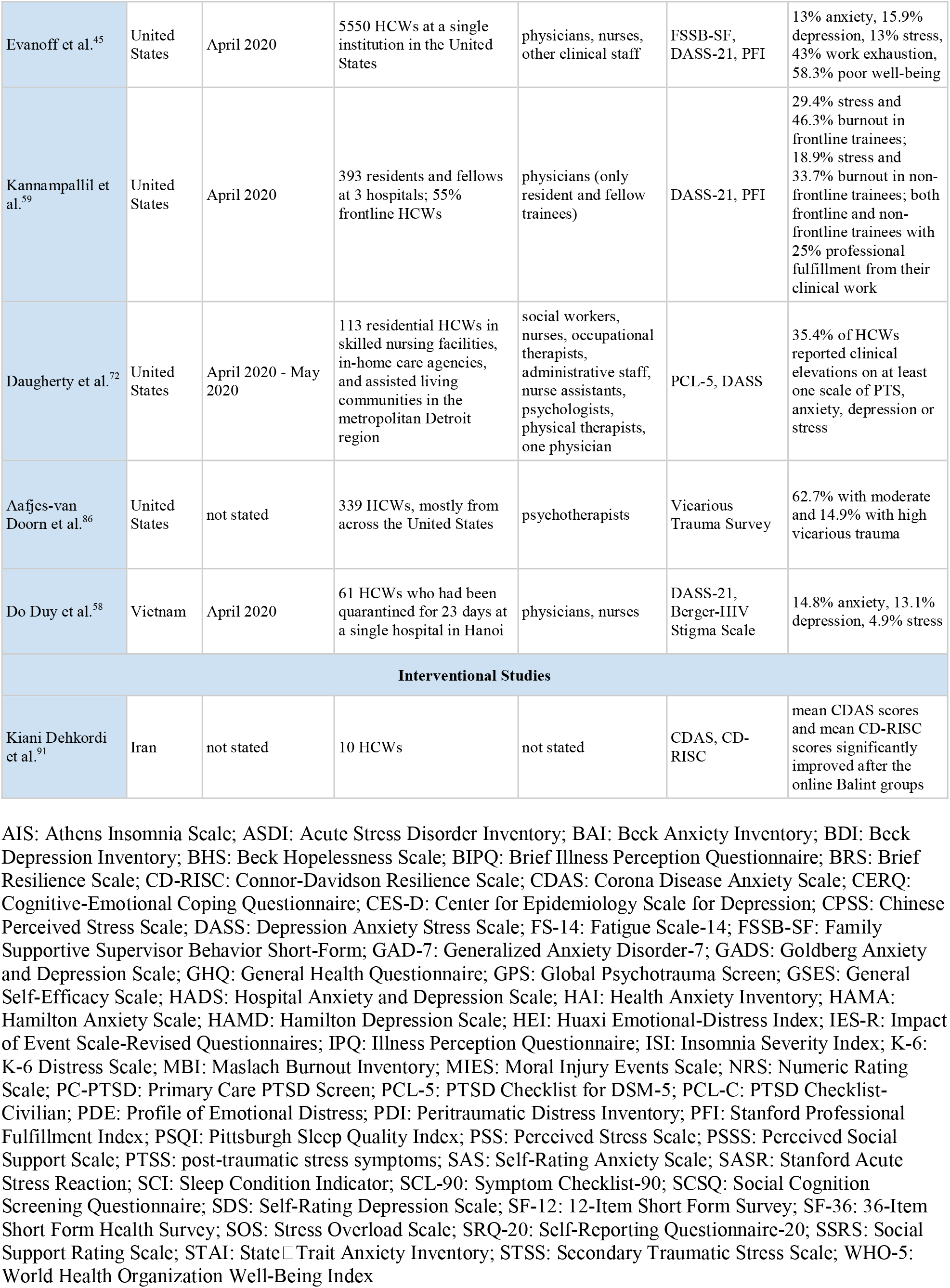
Summary of all included studies.

| Name | Country | Dates of Surveys | Number and Location | Type of HCWs | Outcomes | Findings |
| --- | --- | --- | --- | --- | --- | --- |
| Epidemiologic Studies |  |  |  |  |  |  |
| Xiao et al.<sup>31</sup> | China | January 2020 | 958 HCWs across 26 provinces in China, 65.9% frontline HCWs | physicians, nurses, technicians, other HCWs | PSS-14, HADS | 54.1% anxiety, 57.3% depression |
| Lai et al.<sup>14</sup> | China | January 2020 - February 2020 | 1257 HCWs in 34 hospitals across China, 41.5% frontline HCWs | physicians, nurses | PHQ-9, GAD-7, ISI, IES-R | 44.6% anxiety, 50.4% depression, 34.0% insomnia, 71.5% stress |
| Kang et al.<sup>47</sup> | China | January 2020 - February 2020 | 994 HCWs in Wuhan | physicians, nurses | PHQ-9, GAD-7, ISI, IES-R | 36.9% subthreshold mental health disturbances, 34.4% mild disturbances, 22.4% moderate disturbances, 6.2% severe disturbances |
| Xiao et al.<sup>71</sup> | China | January 2020 - February 2020 | 180 HCWs from several provinces in China, all frontline HCWs | physicians, nurses | SSRS, SAS, GSES, SASR, PSQI | levels of social support for HCWs was significantly associated with self-efficacy and sleep quality, and negatively associated with degree of anxiety and stress |
| Wang et al.<sup>74</sup> | China | January 2020 - February 2020 | 123 HCWs at a single pediatric institution in Wuhan | physicians, nurses | PSQI, SAS, SDS | 7% anxiety, 25% depression, 38% insomnia |
| Xu et al.<sup>87</sup> | China | January 2020 - February 2020 | 60 HCWs in outbreak period and 60 HCWs in non-epidemic outbreak period at a single institution in China | not stated | Anxiety Scale, Depression Score, Dream Anxiety Score, SF-36 | anxiety, depression, dream anxiety, and SF-36 scores were higher in the outbreak period than the non-outbreak period |
| Sun et al.<sup>57</sup> | China | January 2020 - February 2020 | 442 HCWs across China, 52% frontline HCWs | physicians, nurses, administrative and logistics staff, and other | IES-R | median IES-R score of HCWs was at a mild level |
| Xing et al.<sup>49</sup> | China | January 2020 - February 2020 | 548 HCWs across 8 Chinese provinces | physicians, nurses | SCL-90 | 32.7% higher scores on SCL-90 than the national normal score |
| Lin et al.<sup>15</sup> | China | January 2020 - February 2020 | 2316 HCWs at hospitals in Hubei Province, 38% frontline HCWs | physicians, nurses | PHQ-9, GAD-7, ISI, IES-R | 41.1% anxiety, 46.9% depression, 32% insomnia, 69.1% stress |
| Zhang et al.<sup>80</sup> | China | January 2020 - February 2020 | 1563 HCWs from across China | physicians, nurses, administrators, other HCWs | PHQ-9, GAD-7, ISI, IES-R | 44.7% anxiety, 50.7% depression, 36.1% insomnia, 73.4% stress |
| Mo et al.<sup>63</sup> | China | February 2020 | 180 HCWs in Guangxi Province | nurses | SOS, SAS | 39.9% stress overload |
| Huang et al.<sup>77</sup> | China | February 2020 | 7236 citizens across China, 31% HCWs | not stated | GAD-7, CES-D, PSQI | 35.1% anxiety, 20.1% depression, 18.2% insomnia |
| Liu et al.<sup>46</sup> | China | February 2020 | 512 HCWs across China, 32% frontline HCWs | physicians, nurses, administrative workers | SAS | 12.5% anxiety |
| Wu et al.<sup>52</sup> | China | February 2020 | 60 frontline HCWs at a designated COVID-19 hospital and 60 frontline HCWs at a non-designated hospital | not stated | SCL-90, SDS, SAS, PSQI, PCL-C | 61.7% moderate insomnia, 26.7% severe insomnia |
| Yin et al.<sup>8</sup> | China | February 2020 | 371 HCWs across China | physicians, nurses, technicians | PCL-5, PSQI | 3.8% PTS |
| Hou et al.<sup>9</sup> | China | February 2020 | 1472 HCWs in Jiangsu Province | not stated | SSRS, CD-RISC, SCL-90 | 7% psychological abnormality |
| Que et al.<sup>12</sup> | China | February 2020 | 2285 HCWs across 28 provinces in China | physicians, nurses, technicians and public health professionals | GAD-7, PHQ-9, ISI | 46.0% anxiety, 44.4% depression, 28.8% insomnia, 56.6% overall psychological problems |
| Li et al.<sup>55</sup> | China | February 2020 | 948 HCWs, 219 in Wuhan and 729 in Ningbo | physicians, nurses | AIS, SRQ-20 | 58.9% insomnia and 13.2% general psychological problems in Wuhan; 25.0% insomnia and 8.6% general psychological problems in Ningbo |
| Huang et al.<sup>21</sup> | China | February 2020 | 377 HCWs at 32 public hospitals in Sichuan Province | nurses, technicians | SAS, CD-RISC | 23.4% anxiety |
| Tu et al.<sup>73</sup> | China | February 2020 | 100 frontline nurses at a single institution in Wuhan, China | nurses | PSQI, GAD-7, PHQ-9 | 40% anxiety, 46% depression, 60% insomnia |
| Du et al.<sup>32</sup> | China | February 2020 | 200 frontline HCWs from 2 Wuhan-based hospitals and 110 HCWs in the outreach team deployed to Wuhan from 2 outside hospitals. | physicians, nurses, support staff | PSS, BDI-II, BAI | 20.1% anxiety, 12.7% depression, 59.0% stress |
| Cheng et al.<sup>61</sup> | China | February 2020 | 534 pediatric HCWs at 9 hospitals in Jiangsu Province | physicians, nurses | SAS, PSQI | 14.0% anxiety |
| Liu et al.<sup>62</sup> | China | February 2020 | 2031 pediatric HCWs from 29 provinces across China, 36.5% frontline | physicians, nurses | DASS | 18.3% anxiety, 14.8% depression, 10.0% stress |
| Cai et al.<sup>5</sup> | China | February 2020 | 1173 frontline HCWs and 1173 age- and sex-matched non-frontline HCWs across China | physicians, nurses, other HCWs | BAI, ISI, PHQ-9 | 52.6% any mental problem, 15.7% anxiety, 14.3% depression, and 47.8% insomnia in frontline HCWs; 34.0% any mental problem, 7.4% anxiety, 10.1% depression, and 29.1% insomnia in non-frontline HCWs |
| Yang et al.<sup>22</sup> | China | February 2020 | 449 otolaryngology HCWs in Hubei Province | physicians, nurses | SAS | 29.2% anxiety |
| Zhu et al.<sup>33</sup> | China | February 2020 | 5062 HCWs at a single institution in Wuhan, China; 34.3% frontline | physicians, nurses, technicians | PHQ-9, GAD-7, IES-R | 24.1% anxiety, 13.5% depression, 29.8% stress |
| Li et al.<sup>34</sup> | China | February 2020 | 4369 women HCWs at a single institution in Wuhan, China | physicians, nurses, technicians | PHQ-9, GAD-7, IES-R | 25.2% anxiety, 14.2% depression, 31.6% stress |
| Li et al.<sup>85</sup> | China | February 2020 | 214 citizens and 526 nursing HCWs, 44% frontline nurses | nurses | Vicarious Traumatization Questionnaire | vicarious traumatization scores of the general public were similar to those of non-frontline nurses but significantly higher than those of frontline nurses |
| Liang et al.<sup>54</sup> | China | February 2020 | 59 HCWs at a single institution in Guangzhou | physicians, nurses | SDS, SAS | no significant differences in anxiety or depression scores between staff in COVID-associated departments versus other departments |
| Lu et al.<sup>13</sup> | China | February 2020 | 2299 HCWs across Fujian Province | physicians, nurses, administrative staff | NRS, HAMA, HAMD | 70.6% fear, 25.5% anxiety, and 12.1% depression in medical staff; 58.4% fear, 18.7% anxiety, and 8.2% depression in administrative staff |
| Zhang et al.<sup>36</sup> | China | February 2020 - March 2020 | 2182 medical and nonmedical HCWs across China | physicians, nurses, other nonmedical HCWs | PHQ-4, SCL-90-R, ISI | 13.0% anxiety, 12.2% depression, 1.6% somatization, 5.3% obsessive-compulsive symptoms, 38.4% insomnia in medical HCWs; 8.5% anxiety, 9.5% depression, 0.4% somatization, 2.2% obsessive-compulsive symptoms, 30.5% insomnia in nonmedical HCWs |
| Zhu et al.<sup>56</sup> | China | February 2020 - March 2020 | 1443 citizens under quarantine (22% HCWs) and 836 citizens not under quarantine (64% HCWs) | not stated | PHQ-9, GAD-7, SRQ-20 | no significant differences in any of the mental health instruments between cohorts who were quarantined and were not quarantined |
| Si et al.<sup>23</sup> | China | February 2020 - March 2020 | 863 HCWs from 7 provinces in China | physicians, nurses, other HCWs | IES-R, DASS, PSSS, SCSQ | 13.9% anxiety, 13.6% depression, 8.6% stress, 40.2% PTS symptoms |
| Wu et al.<sup>83</sup> | China | March 2020 | 190 HCWs at a single institution in Wuhan, 50% frontline HCWs | physicians, nurses | MBI | 39% burnout in non-frontline HCWs, 13% burnout in frontline HCWs |
| Dong et al.<sup>35</sup> | China | March 2020 | 4618 HCWs from 33 hospitals in Sichuan and Yunnan provinces | physicians, nurses, technicians, administrators | HEI | 24.2% mental health issues |
| Wang et al.<sup>78</sup> | China | March 2020 | 1514 HCWs and 487 non-HCWs in Hubei Province | physicians, nurses | PSQI, HADS | 22.6% anxiety, 35% depression, 61.6% insomnia |
| Zhan et al.<sup>79</sup> | China | March 2020 | 1794 nursing HCWs at four hospitals in Wuhan, 26.5% frontline HCWs | nurses, advanced practice providers | AIS, FS-14, CPSS | 33.9% fatigue, 44.0% stress, 52.8% insomnia |
| Zhan et al.<sup>28</sup> | China | March 2020 | 2667 frontline nurses at 7 hospitals in Wuhan | nurses | FS-14, GAD-7, PHQ-9, CPSS | 39.8% anxiety, 54.7% depression, 62.0% stress, 35.1% fatigue |
| Lam et al.<sup>50</sup> | China | March 2020 - April 2020 | 932 HCWs at 2 provinces in China and Hong Kong | physicians, nurses, other HCWs | PHQ-9 | 50.4% depression in Hong Kong, 15.1% in Hubei, and 12.9% in Guangdong |
| Chen et al.<sup>88</sup> | China | not stated | 92 nurses at T1 and 86 nurses at T2 at a single institution in Wuhan | nurses | SRQ-20 | 26.1% mental health issues at T1 and 9.3% mental health issues at T2 |
| Fan et al.<sup>29</sup> | China | not stated | 25 frontline transdisciplinary nurses (TNs), who did not have experience in respiratory medicine, infectious disease, or emergency and intensive medicine, and 19 frontline non-TNs who did have these experiences, in Hubei Province | nurses | PSS, PSSS | compared with non-TNs, TNs showed higher levels of perceived stress and less perceived social support |
| Cai et al.<sup>64</sup> | China | not stated | 1521 HCWs across hospitals in Jiangsu Province | not stated | SCL-90, CD-RISC, SSRS | 14.1% psychological abnormality |
| Chen et al.<sup>60</sup> | China | not stated | 105 pediatric HCWs in Guiyang |  | SAS, SDS | 18.1% anxiety, 29.5% depression; mean SAS and SDS scores were significantly higher than general population norms |
| Pedrozo-Pupo et al.<sup>89</sup> | Colombia | March 2020 | 407 citizens across Columbia, 44% HCWs | not stated | PSS-10 | 14.3% perceived stress, which was similar between HCWs and other non-HCW professionals |
| Chen et al.<sup>68</sup> | Ecuador | April 2020 - May 2020 | 252 HCWs at 54 health care facilities in 13 provinces in Ecuador | HCWs in hospitals, clinics, emergency response units, medical wards, nursing homes, dental clinics, and pharmacies | GAD-7 | 28.2% anxiety, 32.5% distress |
| Zerbini et al.<sup>27</sup> | Germany | March 2020 - April 2020 | 111 HCWs at a single institution in Germany | physicians, nurses | PHQ-9, MBI | fear of being infected with COVID-19 and work stress were both correlated with increased burnout, depression, anxiety and perceived stress |
| Varshney et al.<sup>90</sup> | India | March 2020 | 653 citizens across India, 22% HCWs | not stated | IES-R | HCWs did not have a significant difference in psychological distress compared to non-HCWs |
| Khanna et al.<sup>66</sup> | India | April 2020 | 2355 ophthalmology attending physicians and resident physicians across India | physicians | PHQ-9 | 32.6% depression |
| Vahedian-Azimi et al.<sup>6</sup> | Iran | February 2020 - March 2020 | 886 citizens, 25% HCWs and 23% medical students, with all HCWs and medical students from a single institution | physicians, nurses, medical students | DASS-21 | 97.3% severe anxiety, 49.7% severe depression, 55.7% severe and extremely severe stress |
| Zhang et al.<sup>37</sup> | Iran | April 2020 | 304 HCWs across Iran | physicians, nurses | SF-12, K-6, PHQ-4 | 28.0% anxiety, 20.6% depression, 20.1% distress |
| Corbett et al.<sup>38</sup> | Ireland | April 2020 - May 2020 | 240 HCWs at a single institution | physicians, nurses, laboratory staff, administrative staff, midwives | PHQ-9, GAD-7 | 21.0% anxiety, 20.3% depression |
| Rossi et al.<sup>39</sup> | Italy | March 2020 | 1379 HCWs across Italy, 53% frontline HCWs | physicians, nurses, ancillary health care workers | PHQ-9, GAD-7, ISI, PSS, GPS | 19.8% anxiety, 24.7% depression, 21.9% stress, 8.3% insomnia, 49.4% PTS |
| Di Tella et al.<sup>40</sup> | Italy | March 2020 - April 2020 | 145 HCWs in the Piedmont Region in Italy | physicians, nurses | quality of life and health-related Visual Analogue Scales, STAI Y1, BDI-II, PCL-5 | HCWs working in COVID-19 wards reported higher levels of depression and PTS symptoms than those working in other healthcare units |
| Magnavita et al.<sup>48</sup> | Italy | March 2020 - April 2020 | 595 HCWs: 82 HCWs with positive COVID-19 testing, 152 HCWs exposed to COVID-19, and 361 control HCWs, at 2 hospitals in Italy | physicians, nurses, technicians, clerks, other HCWs | GADS, SCI, procedural justice in the Colquitt questionnaire, occupational stress in the Siegrist's effort/reward imbalance model | 16.6% anxiety, 20.3% depression |
| De Sio et al.<sup>16</sup> | Italy | April 2020 | 695 HCWs across Italy | physicians | GHQ-12, WHO-5 | 93.8% psychological distress and 58.9% poor well-being in the most affected areas |
| Barello et al.<sup>82</sup> | Italy | not stated | 376 HCWs, all frontline HCWs | physicians, nurses, other HCWs | MBI | 37% emotional exhaustion, 24.7% depersonalization, 53.2% personal gratification |
| Asaoka et al.<sup>81</sup> | Japan | March 2020 - April 2020 | 331 medical and psychiatric disaster HCWs across Japan | physicians, nurses, other HCWs | IES-R, PDI, Stress-Related Questions | anxiety about infection, exhaustion, peritraumatic distress, and medical (rather than psychiatric) HCWs were associated with PTS symptoms |
| Badahdah et al.<sup>17</sup> | Oman | April 2020 | 509 HCWs at 10 health care facilities across Oman; 29% frontline HCWs | physicians, nurses | GAD-7, PSS-10, WHO-5 | 25.9% anxiety, 56.4% stress, 43.4% poor well-being |
| Shahzad et al.<sup>51</sup> | Pakistan | March 2020 - May 2020 | 345 HCWs from hospitals in Punjab | frontline paramedics | BIPQ | perceived threat of COVID-19 infection was positively associated with anxiety, depression, and emotional exhaustion |
| Sandesh et al.<sup>7</sup> | Pakistan | May 2020 | 112 HCWs at hospitals in Karachi | not stated | DASS-21 | 85.7% anxiety, 72.3% depression, 90.1% stress |
| Osama et al.<sup>84</sup> | Pakistan | not stated | 112 surgery residents at a single institution in Pakistan | physicians | MBI | burnout scores were significantly lower during the pandemic compared to before the pandemic among surgical residents |
| Maciaszek et al.<sup>41</sup> | Poland | March 2020 - April 2020 | 1216 HCWs and 823 non-HCWs across Poland | physicians, nurses, paramedics, allied healthcare workers (pharmacists, physiotherapists, occupational therapists, and psychologists), technicians and administrators | GHQ-28 | 60.8% psychopathology among HCWs, 48.0% psychopathology among non-HCWs |
| Szepietowski et al.<sup>67</sup> | Poland | not stated | 123 HCWs at a single institution in Poland | physicians, nurses | PHQ-9, GAD-7, HADS | 29.2%-45% anxiety, 14.2-20% depression |
| Man et al.<sup>53</sup> | Romania | not stated | 115 HCWs at a single institution in Romania, 58% frontline HCWs | physicians, nurses, medical assistants | IPQ, PSS-10, PDE, CERQ | no difference in mental health outcomes between frontline and non-frontline HCWs |
| Temsah et al.<sup>70</sup> | Saudi Arabia | February 2020 | 582 HCWs at a single institution in Saudi Arabia | physicians, nurses, midwives, ancillary health care workers | GAD-7 | 31.8% moderate or higher anxiety; 41.5% of HCWs more stressed from COVID-19 than previous MERS-CoV epidemic |
| Al-Hanawi et al.<sup>18</sup> | Saudi Arabia | May 2020 | 3036 citizens of Saudi Arabia, 31.4% HCWs and 14.8% frontline HCWs, across 13 administrative regions in Saudi Arabia | not stated | COVID-19 Peritraumatic Distress Index | 39.9% severe distress in HCWs, significantly higher than 7.2% severe distress in the general public |
| Stojanov et al.<sup>19</sup> | Serbia | not stated | 118 frontline HCWs and 83 non-frontline HCWs at a single institution in Serbia | physicians, nurses | GAD-7, SDS, SF-36, PSQI | frontline HCWs had worse anxiety, depression, and insomnia than non-frontline HCWs |
| Tan et al.<sup>10</sup> | Singapore | February 2020 - March 2020 | 470 HCWs at 2 institutions in Singapore | physicians, nurses, allied health professionals, pharmacists, technicians, administrators, clerical staff, maintenance workers | DASS-21, IES-R | 14.5% anxiety, 8.9% depression, 6.6% stress, 7.7% PTSD |
| Chew et al.<sup>11</sup> | Singapore and India | February 2020 - April 2020 | 906 HCWs at 5 hospitals in Singapore and India, majority from Singapore | physicians, nurses, allied health professionals | DASS-21, IES-R | 8.7% anxiety, 5.3% depression, 2.2% stress, 3.8% psychological distress; 7.4% PTS |
| Wong et al.<sup>65</sup> | Singapore | March 2020 | 62 HCWs in an outpatient musculoskeletal clinic at a single institution in Singapore | physicians, nurses, technicians, health care assistants, patient service associates | Caregiver Strain Index | 51.6% strain |
| Teo et al.<sup>75</sup> | Singapore | May 2020 - June 2020 | 122 laboratory HCWs at 3 hospitals in Singapore | laboratory technicians | GAD-7, SDS, fear NRS | 53.4% anxiety, 62.1% depression, 55.3% fear |
| Yang et al.<sup>69</sup> | South Korea | April 2020 | 65 physical therapists at 3 university hospitals in South Korea | physical therapists | PHQ-9, GAD-7 | 32.3% anxiety, 18.5% depression |
| García-Fernández et al.<sup>24</sup> | Spain | March 2020 - April 2020 | 781 HCWs and 1006 non-HCWs across Spain | physicians, nurses, other HCWs | HAMA, BDI, ASDI | no difference in anxiety or depression between HCWs and non-HCWs, but acute stress was higher in HCWs |
| Luceño-Moreno et al.<sup>42</sup> | Spain | April 2020 | 1422 HCWs across Spain | physicians, nurses, medical assistants, other HCWs | IES-R, HADS, MBI, BRS | 58.6% anxiety, 46% depression, 56.6% PTS, 41.1% burnout |
| Yildirim et al.<sup>43</sup> | Turkey | March 2020 - April 2020 | 270 HCWs at a single institution in Turkey | physicians, nurses, technicians, other HCWs | HAI, BDI | HCWs with direct contact with COVID-19 patients had worse anxiety and depression than HCWs without contact |
| Cici et al.<sup>30</sup> | Turkey | March 2020 - April 2020 | 461 nursing students at a single institution in Turkey | nurses | STAI-I, BAI | 51% anxiety |
| Hacimusalar et al.<sup>25</sup> | Turkey | April 2020 | 1121 HCWs and 1035 non-HCWs across Turkey | physicians, nurses, other HCWs | STAI, BHS | anxiety and hopelessness scores were higher in HCWs than non-HCWs |
| Arpacioglu et al.<sup>20</sup> | Turkey | May 2020 | 251 HCWs (43% frontline) and 312 non-HCWs across Turkey | physicians, nurses, other HCWs | PHQ-4, STSS | frontline HCWs had significant higher anxiety, depression and secondary traumatization scores than non-frontline HCWs or non-HCWs |
| Hines et al.<sup>76</sup> | United States | March 2020 - April 2020 | 219 attendings, residents and fellows at a single institution in Baltimore, Maryland | physicians | IES-R, MIES | increased inpatient time, workplace stress, and insomnia were associated with PTS symptoms; increased inpatient time and insomnia were associated with moral injury |
| Shechter et al.<sup>26</sup> | United States | April 2020 | 657 HCWs at a single institution in New York City | physicians, advanced practice providers, nurses | PC-PTSD, PHQ-2, GAD-2, PSQI, ISI | 33% anxiety, 48% depression, 57% stress, 75% insomnia |
| Civantos et al.<sup>44</sup> | United States | April 2020 | 349 otolaryngology resident and attending physicians across the United States | physicians | Mini-Z Burnout Assessment, GAD-7, IES-R, PHQ-2 | 47.9% anxiety, 10.6% depression, 60.2% distress, 21.8% burnout, 27.5% PTS |
| Evanoff et al.<sup>45</sup> | United States | April 2020 | 5550 HCWs at a single institution in the United States | physicians, nurses, other clinical staff | FSSB-SF, DASS-21, PFI | 13% anxiety, 15.9% depression, 13% stress, 43% work exhaustion, 58.3% poor well-being |
| Kannampallil et al.<sup>59</sup> | United States | April 2020 | 393 residents and fellows at 3 hospitals; 55% frontline HCWs | physicians (only resident and fellow trainees) | DASS-21, PFI | 29.4% stress and 46.3% burnout in frontline trainees; 18.9% stress and 33.7% burnout in non-frontline trainees; both frontline and non-frontline trainees with 25% professional fulfillment from their clinical work |
| Daugherty et al.<sup>72</sup> | United States | April 2020 - May 2020 | 113 residential HCWs in skilled nursing facilities, in-home care agencies, and assisted living communities in the metropolitan Detroit region | social workers, nurses, occupational therapists, administrative staff, nurse assistants, psychologists, physical therapists, one physician | PCL-5, DASS | 35.4% of HCWs reported clinical elevations on at least one scale of PTS, anxiety, depression or stress |
| Aafjes-van Doorn et al.<sup>86</sup> | United States | not stated | 339 HCWs, mostly from across the United States | psychotherapists | Vicarious Trauma Survey | 62.7% with moderate and 14.9% with high vicarious trauma |
| Do Duy et al.<sup>58</sup> | Vietnam | April 2020 | 61 HCWs who had been quarantined for 23 days at a single hospital in Hanoi | physicians, nurses | DASS-21, Berger-HIV Stigma Scale | 14.8% anxiety, 13.1% depression, 4.9% stress |
| **Interventional Studies** |  |  |  |  |  |  |
| Kiani Dehkordi et al.<sup>91</sup> | Iran | not stated | 10 HCWs | not stated | CDAS, CD-RISC | mean CDAS scores and mean CD-RISC scores significantly improved after the online Balint groups |

**Figure 1.**
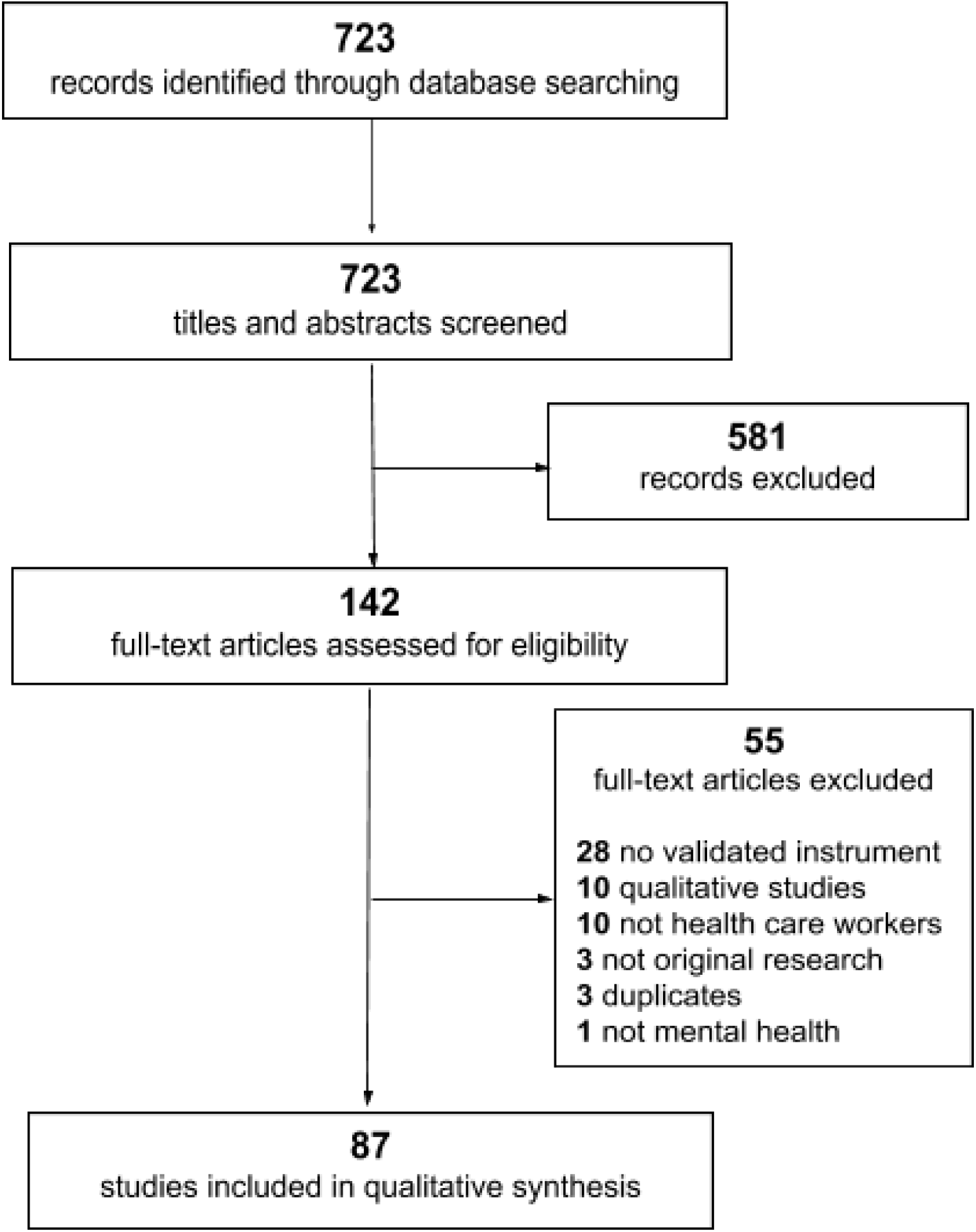
PRISMA flow diagram for rapid reviews.

As shown in **Table 1**, the majority of studies were from China (n = 43, 49%), followed by the United States (n = 7), Italy (n = 5), Singapore (n = 3), Turkey (n = 4), Iran (n = 3), Pakistan (n = 3), India (n = 2), Poland (n = 2), Saudi Arabia (n = 2), Spain (n = 2), Colombia (n = 1), Ecuador (n = 1), Germany (n = 1), Ireland (n = 1), Japan (n = 1), Oman (n = 1), Romania (n = 1), Serbia (n = 1), Singapore and India (n = 1), South Korea (n = 1), and Vietnam (n = 1). The majority of studies occurred early in the pandemic, with 10 studies beginning in January, 27 in February, 20 in March, 15 in April, and 4 in May. Nearly all of the epidemiologic studies were cross-sectional survey-based studies, with the exception of one case-control study.^5^

As displayed in **Table 1**, there was considerable heterogeneity in the mental health outcomes studied and the types of quantitative mental health instruments used. Prevalence of mental health outcomes also varied widely, from 7.0-97.3% anxiety, 10.6-62.1% depression, 2.2-93.8% stress, 3.8-56.6% post traumatic stress (PTS), 8.3-88.4% insomnia, and 21.8-46.3% burnout. These outcomes also varied by country and over time, with very high levels of mental illness in Iran in February^6^ and Pakistan in May,^7^ compared to very low levels in China^8,9^ and Singapore^10,11^ in February.

### Risk factors for adverse mental health outcomes

The majority of studies were epidemiologic and assessed risk factors for adverse mental health or psychological outcomes for HCWs during the COVID-19 pandemic. Risk factors are summarized in **Table 2** and are categorized as follows.

**Table 2.** Summary of risk factors and protective factors for mental health in health care workers.

| **Risk Factors** |
| --- |
| Frontline HCWs |
| HCWs with COVID-19 infection and HCWs exposed to COVID-19 |
| Nurses |
| Nursing students |
| Increased work-hours |
| Increased night shifts |
| Lack of intensive care or public health emergency experience |
| Women |
| Direct contact with COVID-19 patients |
| Working in COVID-19 designated hospitals or isolation wards |
| Working in a geographic region with more COVID-19 cases |
| Personal fear of infection and perceived threat of contagion |
| Friends or family with COVID-19 infection |
| Colleagues hospitalized or died of COVID-19 |
| Trainee physicians, especially frontline trainees |
| Medical students |
| Pediatric HCWs |
| Lack of access to personal protective equipment |
| Poor supervisory support |
| Lack of psychological support |
| Younger HCWs |
| Comorbid medical or mental illness |
| Low psychological resilience |
| Increased attention paid to negative information about the pandemic |
| Increased social media use |
| Lack of knowledge about COVID-19 |
| Presence of physical symptoms |
| Loneliness |
| Presence of children in the home |
| Lack of family support |
| Being an only child |
| Poor sleep quality |
| Presence of burnout |
| **Protective Factors** |
| Social support |
| Personal resilience |
| Greater institutional and administrative support |
| Access and satisfaction with personal protective equipment |
| Access to psychological support |
| Exercise |
| Higher education level |
| Physicians |
| More knowledge about COVID-19 |

#### Frontline workers

Frontline HCWs had worse mental health outcomes than non-frontline HCWs in multiple studies, including increased rates and severity of anxiety, depression, fear, and distress.^12,13,14,15,16,17,18,19,20^ The only case-control study in this review compared 1173 frontline HCWs with 1173 age- and sex-matched non-frontline HCWs across China and found that frontline HCWs had significantly higher rates of any mental health issue than non-frontline HCWs, at 52.6% versus 34.0%, but no significant difference in suicidal ideation, help-seeking, or mental health treatment.^5^

#### Nurses

Nurses experienced poor mental health outcomes compared to physicians and other HCWs,^14,21,22,23,24,25,26^ with several studies identifying specific subsets of the nursing profession at risk for adverse mental health outcomes. Nurses in COVID-19 wards had higher self-reported rates of stress, exhaustion, and depression and lower job fulfillment than nurses in non-COVID wards,^27^ nurses with more work-hours and more night shifts experienced more fatigue,^28^ and nurses without intensive care unit (ICU) experience deployed to COVID-19 ICUs experienced more stress.^29^ Physical and mental fatigue have been associated with anxiety, depression and stress in nurses as well.^28^ One study in Turkey surveyed nursing students before and after the outbreak and found that over half of the nursing students reported anxiety, and anxiety was associated with negative perspectives on the nursing profession as well as unwillingness to practice the profession of nursing in the future.^30^

#### Women

Many studies found that women experience worse mental health outcomes on multiple instruments as compared to men.^6,8,14,17,31,32,33,34,35,36,37,38,39,40,41,42,43,44,45^

#### COVID exposure and infection

Direct contact with COVID-19 patients was associated with poor mental health outcomes,^8,13,31,35,36,43,45,46,47,48^ as well as personal fear of infection and perceived threat of contagion.^13,21,22,27,32,37,49,50,51^ These adverse outcomes were also seen in HCWs with friends or family with COVID-19 infection^22,32,33,34,49^ and in HCWs with colleagues who were hospitalized or died of COVID-19.^39^ Further, working in COVID-19 designated hospitals compared to non-designated hospitals^25,49,52^ and working in COVID-19 specific isolation wards^13,22,40,45^ were associated with poorer mental health outcomes. However, smaller single-institution studies in Romania^53^ and in Guangzhou, China,^54^ where there were fewer COVID-19 cases, did not find differences in mental health outcomes between HCWs working in dedicated versus non-dedicated wards. In this same vein, geographic regions with higher rates of COVID-19 cases, such as Wuhan and Hubei province, had worse mental health outcomes than other less affected areas.^9,14,32,46,49,50,55^ This finding was also demonstrated in Italy^16^ and the United States.^44^ One study of 595 HCWs in Italy compared mental health outcomes between HCWs with positive COVID-19 testing, HCWs with COVID-19 exposure, and control HCWs, and found fourfold worse anxiety and depression in COVID-infected HCWs.^48^

#### Quarantine

One large study in China of 1443 Chinese citizens under quarantine (22% HCWs) and 836 Chinese citizens not under quarantine (64% HCWs) did not find a difference in anxiety, depression or distress,^56^ although PTS was found to be more severe in quarantined HCWs in a separate study.^57^ A single-institution study of quarantined HCWs in Vietnam found evidence of social stigma, negative self-image, and guilt towards family and friends.^58^

#### Trainees

Several studies assessed mental health in trainee physicians, finding more depression^24^ and loneliness^26^ in trainees compared to other HCWs. A large multicenter survey of trainees in the United States found that frontline trainees have worse burnout and stress, but similar anxiety and depression, to non-frontline trainees, and that trainees who were COVID-19 exposed, women, and unmarried also have worse outcomes.^59^ Medical students have also been studied, with one single-institution study in Iran of 886 Iranian citizens, which included 207 (23%) medical students, finding worse mental health in medical students compared to medical staff and the general population.^6^

#### Pediatric HCWs

Several studies assessed pediatric HCWs, revealing that they reported poorer mental health outcomes than the general population.^60^ With respect to risk factors, working in outpatient, emergency and ICU departments^61^ and COVID-19 exposure^60,62^ were associated with worse mental health outcomes. A large multicenter study of 2031 pediatric HCWs from 29 provinces across China found that frontline HCWs, men, physicians, HCWs aged 31-60 years, HCWs with senior job titles, and HCWs who had experience combating similar outbreaks were more likely to have stress, depression and anxiety.^62^

#### Organizational characteristics

Lack of access to adequate personal protective equipment (PPE) was associated with increased anxiety and depression in several studies.^13,21,31,50^ Poor supervisor support was associated with worse mental health in a large single-institution study of 5550 HCWs in the United States.^45^

#### Professional characteristics

Various professional risk factors for poor mental health outcomes were identified, including increased work-hours,^24,42,63^ more than 10 years of working experience,^33,34^ HCWs without public health emergency experience,^64^ hospital-based physicians,^16^ more night shifts,^16^ lower job seniority,^16^ significant work adjustments,^65^ physicians worried about training or professional growth,^66^ and physicians with difficulty meeting living expenses.^66^

#### Personal characteristics

The majority of studies found that younger HCWs had worse mental health outcomes than older HCWs,^17,37,38,39,41,42,43,45,66^ although one study of nurses and health care technicians in radiology departments in China found that older HCWs had increased anxiety.^21^ HCWs with comorbid chronic diseases,^33,34,36,67^ history of mental illness,^33,34^ and increased alcohol consumption^16^ experienced poorer mental health outcomes. Low psychological resilience and worse perceived self-efficacy were associated with worse mental health,^21,32^ and increased attention paid to negative information about the pandemic were also associated with worse outcomes.^12,61^ One multinational study in Singapore and India found that presence of physical symptoms was associated with significantly worse anxiety, depression, stress, and post-traumatic stress disorder (PTSD).^11^ One small study in China found that lack of knowledge about COVID-19 was associated with worse mental health outcomes,^32^ and another small study in Ecuador found that belief in conspiracy theories surrounding the origin of COVID-19 was associated with worse mental health in HCWs.^68^

#### Social supports

Mental health outcomes were poorer in HCWs who were only children,^63^ reported loneliness,^13,16,38^ had children of their own,^34,69^ lived in rural areas,^36^ and felt lack of family support.^12,32,49^

#### Previous epidemic exposure

A single-institution study conducted in Saudi Arabia surveyed HCWs in February prior to any documented COVID cases in the country and found moderate to severe anxiety in over 31.8% of HCWs.^70^ Additionally, 41.5% of HCWs were more stressed about COVID-19 than they had about the previous MERS-CoV SARS outbreak in 2002-2003.

### Protective factors for mental health

Social support was found to be a protective factor for mental health in HCWs in several studies,^9,23,27,51,71^ including one study which found that personal resilience mediated the relationship between social support and mental health.^9^ Further, greater support at work was found to mitigate adverse mental health outcomes in one multicenter study in the United States,^72^ and a large single-institution study in China found that increased perception of institutional and administrative support, as well as satisfaction with PPE, were associated with less stress among HCWs.^33^ Several other studies also found that adequate access to PPE was associated with better mental health outcomes.^37,41^ In one large multicenter study in China, HCWs receiving psychological support were significantly less likely to report adverse mental health outcomes.^15^ HCWs with higher psychological resilience,^21^ more frequent exercise,^28,34^ higher education level,^37,42^ physicians,^42^ more knowledge about COVID-19,^43^ and HCWs working in private hospitals in Iran^37^ were found to have better mental health.

### Sleep quality

Numerous studies in this review specifically assessed sleep and risk factors for poor sleep quality in HCWs. Sleep quality was related to anxiety, depression and PTS in several studies.^19,32,61,73,74,75,76^ Sleep quality was also found to mediate the association between COVID-19 exposure level and PTS symptoms^8^ as well as the association between occupational stress and anxiety.^48^ Improved sleep quality was found in HCWs with adequate social support.^71^ HCWs reported more sleep disturbances than non-HCWs in several studies,^41,77,78^ as well as worse sleep in frontline HCWs compared to non-frontline HCWs^78,79^ and worse sleep in medical HCWs compared to non-medical HCWs.^36^ Risk factors for worse sleep quality include nurses,^26,39^ HCWs working in Wuhan, China,^55^ working in a COVID-designated hospital or COVID isolation unit,^52,80^ fear about infection and uncertainty about epidemic control,^80^ lack of psychological support,^80^ health care assistants,^39^ direct contact with COVID patients,^74^ being an only child,^74^ and having a colleague die due to COVID-19.^39^

### Acute traumatic stress and PTS symptoms

The presence of acute traumatic stress and PTS symptoms in HCWs was assessed in numerous studies in this review. Nonmedical HCWs were found to have increased PTS compared to medical HCWs in one study in Singapore, although the overall prevalence of PTS was much lower at 7.7% compared to other studies.^10^ Risk factors for increased PTS symptoms included working in a COVID-designated hospital or ward,^40,52^ working in a region with more COVID cases,^42^ direct contact with COVID patients,^8^ women,^40,42^ older HCWs,^40,57^ quarantined HCWs,^57^ frontline HCWs,^39^ general practitioners,^39^ having a colleague be hospitalized or die from COVID,^39^ anxiety about COVID infection,^81^ exhaustion,^81^ and working night shifts.^42^ A single-institution study of physicians in the United States found higher PTS symptoms in HCWs with more inpatient shifts, higher perceived workplace stress, and increased sleep difficulties, although the study also found that greater frequency of shift work was protective of PTS.^76^ A multicenter study of 349 otolaryngology physicians in the United States found 27.5% reported moderate to severe PTS.^44^

### Burnout

A large multicenter study of 1422 HCWs in Spain found 41.1% prevalence of burnout, with burnout positively and significantly related to anxiety, depression and PTS symptoms.^42^ Risk factors for burnout identified in this review included women HCWs,^44,82^ nurses,^82^ direct contact with COVID patients,^45^ fear of infection,^27^ increased work stress,^27^ and presence of somatic symptoms.^82^ In a single-institution study in China, burnout was significantly lower in frontline HCWs compared to non-frontline HCWs.^83^ However, in a multicenter study of trainee physicians in the United States, frontline trainees had higher burnout scores than non-frontline trainees.^59^ Burnout scores in trainees varied depending on the specialty studied, with lower burnout scores in surgery residents in a single-center study in Pakistan compared to before the pandemic, likely due to less work-hours,^84^ but higher burnout scores in otolaryngology residents compared to attending physicians in a multicenter study in the United States.^44^

### Secondary traumatic stress and vicarious traumatization

Several studies specifically assessed vicarious traumatization, also known as secondary traumatic stress, in HCWs. One multicenter study in China compared vicarious traumatization scores of 214 non-HCW citizens with 526 nurses and found that the general public and non-frontline nurses had higher vicarious traumatization scores than frontline nurses.^85^ This is contrary to a multicenter study in Turkey, which also compared 312 citizens with 251 HCWs and found worse secondary traumatic stress scores in frontline HCWs compared to non-frontline HCWs and the general public.^20^ This study also found worse secondary traumatic stress in HCWs who were women, early in their careers, living with parents, had a chronic disease, and reported increased social media use.^20^ Another multicenter study of 339 psychotherapy HCWs in the United States found 77.6% of them had at least moderate vicarious trauma, with higher trauma scores in younger and early-career therapists.^86^

### Moral distress and injury

One single-institution study in the United States specifically assessed moral injury in 219 HCWs, specifically trainees and attending physicians.^76^ This study found that a higher proportion of inpatient time and insomnia were associated with higher moral injury, while a supportive workplace environment was associated with less moral injury.

### Longitudinal assessments of mental health

There were two studies which performed longitudinal assessments of mental health, both performed in China. In the first single-institution study, 60 HCWs were surveyed during the outbreak from January to February 2020, and then 60 different HCWs were surveyed after the outbreak in March 2020.^87^ The study found that mental health outcomes were worse during the outbreak compared to the post-outbreak period. A similar single-institution study surveyed 92 nurses after they had worked 7-10 days in COVID isolation wards and compared them to 86 nurses after they had worked 2 months in isolation wards.^88^ This study also found that mental health outcomes were worse earlier during the outbreak, and additionally found that there were different stressors and mental health needs by HCWs during both time periods.

### Comparison between medical HCWs, nonmedical HCWs, and non-HCWs

Studies which compared mental health outcomes of HCWs with non-HCWs generally found that HCWs had worse outcomes than non-HCWs in the general population, including in China,^49^ Poland,^41^ Saudi Arabia,^18^ and Turkey.^25^ However, a study in Colombia found similar rates of stress between HCWs and other professions,^89^ a study in Spain found no difference in anxiety and depression between HCWs and non-HCWs, but did find higher acute stress in HCWs,^24^ and a study in India found no difference in psychological distress in HCWs compared to non-HCWs.^90^ Several studies also compared medical HCWs with non-medical HCWs. A large multicenter study of 2182 HCWs in China found higher anxiety, depression, and insomnia among medical HCWs,^36^ with similar findings of higher fear and anxiety in medical HCWs as compared to non-medical HCWs in another large multicenter study of 2299 HCWs in China.^13^ This is contrary to a multicenter study of 470 HCWs in Singapore which found higher anxiety, stress and PTS among non-medical HCWs compared to medical HCWs.^10^ This was also demonstrated in a single-center study of 240 HCWs in Ireland, which found higher anxiety and depression scores were in administrators compared to medical HCWs.^38^ A multicenter study of 122 laboratory HCWs in Singapore found high rates of depression, anxiety and fear among laboratory technicians, and also found that previous experience handling high-risk specimens during the previous SARS outbreak did not influence mental health outcomes.^75^

### Interventional studies

There was only one interventional study identified in this review that assessed mental health using a validated instrument. The study is a pilot study at a single institution in Iran of 10 HCWs who participated in online Balint groups.^91^ The study found that mean anxiety and resilience scores significantly improved after the online Balint groups.

## Discussion

This rapid review describes risk factors for various adverse mental health outcomes among HCWs during the ongoing COVID-19 pandemic, as well as supportive measures which may mitigate these risks. There is clearly a dire need for effective mental health interventions as new outbreaks develop and HCWs incur repetitive trauma, and further study on other HCW-specific sources of stress, including burnout and moral injury, will be required to comprehensively describe the acute and long-term mental health effects of COVID-19.

The vast majority of studies in this review were cross-sectional survey-based studies that assessed mental health, predominantly anxiety, depression, and acute stress, at a single point in time during the early pandemic. As shown in **Table 1**, there is large between-study heterogeneity in these mental health outcomes, likely due to the wide variety of validated instruments used as well as different definitions or score cutoffs that defined mental illness in each study. There was relatively high prevalence of clinical anxiety and depression among HCWs, with rates appearing to increase in countries other than China as the pandemic continued from March onwards.

Although these cross-sectional studies identified important risk factors, they do not assess how these risk factors or mental health outcomes may change over time. There were only two studies which collected longitudinal data,^87,88^ and both demonstrated poor mental health among HCWs during the main outbreak in China compared to the post-outbreak period. This phenomenon has been previously demonstrated in longitudinal studies during other epidemics,^92,93^ with research from the SARS era finding that frontline HCWs suffered increased stress up to 1 year after the epidemic compared to non-frontline HCWs.^94^ Future longitudinal studies are necessary as the COVID-19 pandemic continues, as the effect of prolonged trauma on HCWs, including repeated cycles of outbreaks and recovery, is not known.

As shown in **Table 2**, there are considerable similarities between risk factors for poor mental health during COVID-19 and previous epidemics such as SARS, including the important role of adequate training, access to PPE, and clear communication in protecting mental health.^92,95^ However, there are also some key differences, as illustrated in this review. Previous research by Chong et al. modeled two distinct psychological phases of HCWs workers during the SARS outbreak in Taiwan: an initial shock and reaction phase, when SARS cases were exponentially growing, and a repair and reorientation phase, when the outbreak was brought under control.^96^ Each phase had distinct psychological findings in HCWs, with anxiety predominating in the initial phase due to feelings of vulnerability, loss of control, high job-related stress, and perceived threat from lack of safeguards and personal protective equipment. The repair and reorientation phase revealed predominant depression and PTS symptomatology, including avoidance, intrusion, and interpersonal difficulty, with many workers considering resigning during this phase. This landmark study demonstrated that, due to different psychopathologies in HCWs at different phases of epidemics, mental health interventions must be comprehensive to address the changing needs of workers as the pandemic progresses. However, studies from the COVID era have demonstrated high prevalence of anxiety in HCWs in Saudi Arabia^70^ and Iran^6^ before the pandemic reached those countries, suggesting that there may be an antecedent phase of anticipatory anxiety or stress in pandemics that may be amenable to prophylactic intervention to prevent worsening mental health. High prevalence of anticipatory anxiety was not universally demonstrated in this review, with much lower rates of pre-pandemic anxiety in Singapore^10,11^ which may be due to their previous experience with the SARS epidemic.

Quarantine increased the risk of PTSD and mental illness during the SARS epidemic,^97,98^ but findings have been more mixed in the COVID-19 literature, possibly due to reduction of quarantine-related stigma for HCWs as widespread community-level quarantine was used during the initial pandemic. Although burnout was not as extensively studied in the literature at the time of the SARS epidemic, studies found worse burnout in frontline HCWs during SARS,^92^ which was redemonstrated in the COVID-19 era in this review. Only a minority of studies specifically studied burnout, as well as other traumatic exposures such as moral injury and vicarious traumatization, which limits conclusions about how those psychological phenomena may affect HCWs. Moral injury in particular suffers from a lack of validated instruments in HCWs, and there are not currently well-described approaches to treating moral injury in HCWs, with further study needed.^99,100^

Access to mental health services and psychological support were found to be protective of adverse mental health outcomes in this review, although there is a suggestion that frontline HCWs may not seek out or receive mental health treatment despite their higher rates of mental illness.^5^ This may represent an unmet need for this at-risk population that is amenable to targeted organizational interventions. Several studies in this review also found that higher personal resilience was protective of mental health, including that resilience may mediate the relationship between psychosocial support and mental health.^9^ The sole interventional study in this review found improved anxiety and personal resilience after exposure to online Balint groups,^91^ also suggesting that mental health is interrelated with resilience. These findings have important implications in designing mental health support programs, as a multipronged approach with organizational-level and individual-level interventions is required.

### Evidence-based mental health frameworks

The present review only identified one interventional study which used a validated mental health instrument to study the effects of a mental health program on HCWs. With few COVID-specific interventional studies to guide mental health support for HCWs, the following evidence-based mental health frameworks, many based on previous epidemics which share similarities to COVID-19, can serve as models to execute and study novel mental health interventions.

#### Psychological First Aid

Psychological First Aid (PFA) was developed by the National Center for PTSD in 2006 after it was recognized that traditional trauma debriefing interventions, such as critical incident stress debriefing, did not promote recovery from traumatic incidents and in fact may increase the risk of PTSD.^101,102^ PFA was based on the observation that most psychological responses to mass trauma are appropriate reactions to distressing situations rather than psychiatric illness, and that social support can mitigate this distress.^103^ As such, PFA involves empowering traumatized persons with safety, promotion of self-efficacy, calmness, social connectedness, and optimism, and is designed to be implemented by HCWs or the general public.^102^ PFA is promoted by multiple mental health organizations as an evidence-based approach to supporting HCWs in crisis, although a systematic review did not find strong evidence of its risks or benefits.^104^ Further, because PFA was designed based on the experience of mass casualty or mass trauma incidents, it is unclear if PFA will be effective for repetitive stressors during a pandemic.^105^ A brief PFA model has been developed in Malaysia and implemented with HCWs, although quantitative data are currently lacking.^106,107^

#### Stress Continuum Model and Stress First Aid

With increasing literature suggesting that traumatic stressors are frequently not due to a single event but are instead longitudinal and cumulative in nature, a related psychological support framework was developed by the United States military and called Stress First Aid (SFA).^108^ Unlike PFA, SFA is embedded in a larger Stress Continuum Model which recognizes that traumatized individuals exist on a spectrum of appropriate reactive distress to frank mental illness, and may move along that spectrum as they are exposed to different stressors.^108^ Further, SFA is specifically designed as a peer-to-peer model, and informed the creation of a COVID-19-specific peer-to-peer Battle Buddies intervention with HCWs that is currently under study.^109^ Although there is little research on the efficacy of these models, the findings in this review that mental health outcomes are extremely variable, that there are myriad risk factors which differ based on type of HCW and other personal or professional characteristics, and that social support mitigates the risk of adverse outcomes, suggests that SFA as a comprehensive psychosocial support model may effectively address the needs of the majority of stressed HCWs.

#### Personal resilience training

As low psychological resilience and perceived self-efficacy were associated with worse mental outcomes in this review, study of interventions that build personal resilience in HCWs are warranted. Several computer-based resilience training modules were developed in Toronto after the SARS epidemic, with low-quality evidence of improvement in mental health with these modules.^110^

#### Organizational resilience and organizational justice

Findings from the SARS epidemic that mental health of HCWs was improved when they were provided with adequate training and protective equipment, organizational support, and clear communication from leadership led to the conceptualization of organizational resilience or organizational justice.^110,111^ These frameworks are considered a critical component of pre-pandemic planning, and involve establishment of crisis protocols that provide for the practical and physical needs of HCWs as well as their mental health needs.^112^ These include equitable access to sleep facilities, childcare, food, and psychological support services, in which leadership ensures that HCWs voices are compassionately heard in all decision-making procedures throughout the pandemic. Further, given that lack of information about COVID-19 and increased attention paid to negative information about the pandemic are associated with worse mental health, timely and supportive crisis communication by leadership is felt to be vital in supporting HCWs.^113^

#### Cognitive behavioral therapy and mindfulness

Various cognitive behavioral therapy (CBT) approaches to supporting the mental health of HCWs have been developed, including the Accelerated Recovery Program,^114^ which has demonstrated efficacy in reducing compassion fatigue, burnout, and secondary traumatization.^115,116^ A systematic review of randomized controlled trials has confirmed that CBT and mindfulness improve stress, anxiety and depression in HCWs^117^ and CBT-informed interventions for HCWs during COVID-19 have been proposed.^105^ Although these interventions are efficacious, further study on how they can be incorporated into larger organizational mental health frameworks is warranted.

### Strengths and limitations

Strengths of this rapid review include a broad initial query for all studies on the mental health of HCWs during the COVID-19 pandemic, as well as only including studies that utilized validated mental health instruments. Further, approximately half of the countries represented in this review were outside of China, which enhances the external validity of the review. Limitations include the intrinsic variability and heterogeneity of outcomes and mental health instruments utilized across studies, wide variation in the sample sizes and inclusion of studies with low power, and utilization of only one database. Interventional studies were poorly represented in this review, and future reviews will be required to capture new interventional studies as they are published.

## Conclusions

Research from previous epidemics as well as during the current COVID-19 pandemic suggests that HCWs experience disproportionately poor mental health outcomes compared to the general population, particularly among HCWs working in frontline and high-risk environments.

Although this review captures a large body of literature that robustly describes cross-sectional data on risk factors for adverse mental health outcomes in HCWs across multiple countries, there is limited research on the longitudinal effects of the present pandemic, although longitudinal studies are currently ongoing.^118^ The use of validated mental health instruments is vital, and development of novel instruments for moral injury, secondary traumatic stress, and vicarious traumatization is warranted to elucidate how these phenomena may be affecting HCWs. Further, there is a dearth of literature on interventions that can quantitatively improve mental health outcomes. The heterogeneity of the literature suggests that mental health interventions for HCWs cannot be one size fits all, and must incorporate multifaceted organizational- and individual-level programs that flexibly address the spectrum of acute and chronic stress to which different HCWs are exposed as the pandemic continues.

## Data Availability

Not applicable.

## Acknowledgements

None to declare.

## References

1. Greenberg, N. (2020). Mental health of health-care workers in the COVID-19 era. Nature Reviews. Nephrology, 16(8), 425–426.

2. Maunder, R. G., Lancee, W. J., Balderson, K. E., Bennett, J. P., Borgundvaag, B., Evans, S., Fernandes, C. M. B., Goldbloom, D. S., Gupta, M., Hunter, J. J., McGillis Hall, L., Nagle, L. M., Pain, C., Peczeniuk, S. S., Raymond, G., Read, N., Rourke, S. B., Steinberg, R. J., Stewart, T. E., … Wasylenki, D. A. (2006). Long-term psychological and occupational effects of providing hospital healthcare during SARS outbreak. Emerging Infectious Diseases, 12(12), 1924–1932.

3. Preti, E., Di Mattei, V., Perego, G., Ferrari, F., Mazzetti, M., Taranto, P., Di Pierro, R., Madeddu, F., & Calati, R. (2020). The psychological impact of epidemic and pandemic outbreaks on healthcare workers: Rapid review of the evidence. Current Psychiatry Reports, 22(8), 43.

4. Williams, R. D., Brundage, J. A., & Williams, E. B. (2020). Moral injury in times of COVID-19. Journal of Health Service Psychology, 46(2), 1–5.

5. Cai, Q., Feng, H., Huang, J., Wang, M., Wang, Q., Lu, X., Xie, Y., Wang, X., Liu, Z., Hou, B., Ouyang, K., Pan, J., Li, Q., Fu, B., Deng, Y., & Liu, Y. (2020). The mental health of frontline and non-frontline medical workers during the coronavirus disease 2019 (COVID- 19) outbreak in China: A case-control study. Journal of Affective Disorders, 275, 210–215.

6. Vahedian-Azimi, A., Moayed, M. S., Rahimibashar, F., Shojaei, S., Ashtari, S., & Pourhoseingholi, M. A. (2020). Comparison of the severity of psychological distress among four groups of an Iranian population regarding COVID-19 pandemic. BMC Psychiatry, 20(1), 402.

7. Sandesh, R., Shahid, W., Dev, K., Mandhan, N., Shankar, P., Shaikh, A., & Rizwan, A. (2020). Impact of COVID-19 on the mental health of healthcare professionals in Pakistan. Cureus, 12(7), e8974.

8. Yin, Q., Sun, Z., Liu, T., Ni, X., Deng, X., Jia, Y., Shang, Z., Zhou, Y., & Liu, W. (2020). Posttraumatic stress symptoms of health care workers during the corona virus disease 2019. Clinical Psychology & Psychotherapy, 27(3), 384–395.

9. Hou, T., Zhang, T., Cai, W., Song, X., Chen, A., Deng, G., & Ni, C. (2020). Social support and mental health among health care workers during Coronavirus Disease 2019 outbreak: A moderated mediation model. PloS One, 15(5), e0233831.

10. Tan, B. Y. Q., Chew, N. W. S., Lee, G. K. H., Jing, M., Goh, Y., Yeo, L. L. L., Zhang, K., Chin, H.-K., Ahmad, A., Khan, F. A., Shanmugam, G. N., Chan, B. P. L., Sunny, S., Chandra, B., Ong, J. J. Y., Paliwal, P. R., Wong, L. Y. H., Sagayanathan, R., Chen, J. T., … Sharma, V. K. (2020). Psychological impact of the COVID-19 pandemic on health care workers in Singapore. Annals of Internal Medicine, 173(4), 317–320.

11. Chew, N. W. S., Lee, G. K. H., Tan, B. Y. Q., Jing, M., Goh, Y., Ngiam, N. J. H., Yeo, L. L. L., Ahmad, A., Ahmed Khan, F., Napolean Shanmugam, G., Sharma, A. K., Komalkumar, R. N., Meenakshi, P. V., Shah, K., Patel, B., Chan, B. P. L., Sunny, S., Chandra, B., Ong, J. J. Y., … Sharma, V. K. (2020). A multinational, multicentre study on the psychological outcomes and associated physical symptoms amongst healthcare workers during COVID-19 outbreak. Brain, Behavior, and Immunity, 88, 559–565.

12. Que, J., Shi, L., Deng, J., Liu, J., Zhang, L., Wu, S., Gong, Y., Huang, W., Yuan, K., Yan, W., Sun, Y., Ran, M., Bao, Y., & Lu, L. (2020). Psychological impact of the COVID-19 pandemic on healthcare workers: a cross-sectional study in China. General Psychiatry, 33(3), e100259.

13. Lu, W., Wang, H., Lin, Y., & Li, L. (2020). Psychological status of medical workforce during the COVID-19 pandemic: A cross-sectional study. Psychiatry Research, 288(112936), 112936.

14. Lai, J., Ma, S., Wang, Y., Cai, Z., Hu, J., Wei, N., Wu, J., Du, H., Chen, T., Li, R., Tan, H., Kang, L., Yao, L., Huang, M., Wang, H., Wang, G., Liu, Z., & Hu, S. (2020). Factors associated with mental health outcomes among health care workers exposed to Coronavirus disease 2019. JAMA Network Open, 3(3), e203976.

15. Lin, K., Yang, B. X., Luo, D., Liu, Q., Ma, S., Huang, R., Lu, W., Majeed, A., Lee, Y., Lui, L. M. W., Mansur, R. B., Nasri, F., Subramaniapillai, M., Rosenblat, J. D., Liu, Z., & McIntyre, R. S. (2020). The mental health effects of COVID-19 on health care providers in China. The American Journal of Psychiatry, 177(7), 635–636.

16. De Sio, S., Buomprisco, G., La Torre, G., Lapteva, E., Perri, R., Greco, E., Mucci, N., & Cedrone, F. (2020). The impact of COVID-19 on doctors’ well-being: results of a web survey during the lockdown in Italy. European Review for Medical and Pharmacological Sciences, 24(14), 7869–7879.

17. Badahdah, A., Khamis, F., Al Mahyijari, N., Al Balushi, M., Al Hatmi, H., Al Salmi, I., Albulushi, Z., & Al Noomani, J. (2020). The mental health of health care workers in Oman during the COVID-19 pandemic. The International Journal of Social Psychiatry, 20764020939596.

18. Al-Hanawi, M. K., Mwale, M. L., Alshareef, N., Qattan, A. M. N., Angawi, K., Almubark, R., & Alsharqi, O. (2020). Psychological distress amongst health workers and the general public during the COVID-19 pandemic in Saudi Arabia. Risk Management and Healthcare Policy, 13, 733–742.

19. Stojanov, J., Malobabic, M., Stanojevic, G., Stevic, M., Milosevic, V., & Stojanov, A. (2020). Quality of sleep and health-related quality of life among health care professionals treating patients with coronavirus disease-19. The International Journal of Social Psychiatry, 20764020942800.

20. Arpacioglu, S., Gurler, M., & Cakiroglu, S. (2020). Secondary traumatization outcomes and associated factors among the health care workers exposed to the COVID-19. The International Journal of Social Psychiatry, 20764020940742.

21. Huang, L., Wang, Y., Liu, J., Ye, P., Chen, X., Xu, H., Qu, H., & Ning, G. (2020). Factors influencing anxiety of health care workers in the radiology department with high exposure risk to COVID-19. Medical Science Monitor: International Medical Journal of Experimental and Clinical Research, 26, e926008.

22. Yang, X., Zhang, Y., Li, S., & Chen, X. (2020). Risk factors for anxiety of otolaryngology healthcare workers in Hubei province fighting coronavirus disease 2019 (COVID-19). Social Psychiatry and Psychiatric Epidemiology.

23. Si, M.-Y., Su, X.-Y., Jiang, Y., Wang, W.-J., Gu, X.-F., Ma, L., Li, J., Zhang, S.-K., Ren, Z.-F., Ren, R., Liu, Y.-L., & Qiao, Y.-L. (2020). Psychological impact of COVID-19 on medical care workers in China. Infectious Diseases of Poverty, 9(1), 113.

24. García-Fernández, L., Romero-Ferreiro, V., López-Roldán, P. D., Padilla, S., Calero-Sierra, I., Monzó-García, M., Pérez-Martín, J., & Rodriguez-Jimenez, R. (2020). Mental health impact of COVID-19 pandemic on Spanish healthcare workers. Psychological Medicine, 1–3.

25. Hacimusalar, Y., Kahve, A. C., Yasar, A. B., & Aydin, M. S. (2020). Anxiety and hopelessness levels in COVID-19 pandemic: A comparative study of healthcare professionals and other community sample in Turkey. Journal of Psychiatric Research, 129, 181–188.

26. Shechter, A., Diaz, F., Moise, N., Anstey, D. E., Ye, S., Agarwal, S., Birk, J. L., Brodie, D., Cannone, D. E., Chang, B., Claassen, J., Cornelius, T., Derby, L., Dong, M., Givens, R. C., Hochman, B., Homma, S., Kronish, I. M., Lee, S. A. J., … Abdalla, M. (2020). Psychological distress, coping behaviors, and preferences for support among New York healthcare workers during the COVID-19 pandemic. General Hospital Psychiatry, 66, 1–8.

27. Zerbini, G., Ebigbo, A., Reicherts, P., Kunz, M., & Messman, H. (2020). Psychosocial burden of healthcare professionals in times of COVID-19 - a survey conducted at the University Hospital Augsburg. German Medical Science: GMS e-Journal, 18, Doc05.

28. Zhan, Y.-X., Zhao, S.-Y., Yuan, J., Liu, H., Liu, Y.-F., Gui, L.-L., Zheng, H., Zhou, Y.-M., Qiu, L.-H., Chen, J.-H., Yu, J.-H., & Li, S.-Y. (2020). Prevalence and influencing factors on fatigue of first-line nurses combating with COVID-19 in China: A descriptive cross-sectional study. Current Medical Science, 40(4), 625–635.

29. Fan, J., Hu, K., Li, X., Jiang, Y., Zhou, X., Gou, X., & Li, X. (2020). A qualitative study of the vocational and psychological perceptions and issues of transdisciplinary nurses during the COVID-19 outbreak. Aging, 12(13), 12479–12492.

30. Cici, R., & Yilmazel, G. (2020). Determination of anxiety levels and perspectives on the nursing profession among candidate nurses with relation to the COVID-19 pandemic. Perspectives in Psychiatric Care, ppc.12601. https://doi.org/10.1111/ppc.12601

31. Xiao, X., Zhu, X., Fu, S., Hu, Y., Li, X., & Xiao, J. (2020). Psychological impact of healthcare workers in China during COVID-19 pneumonia epidemic: A multi-center cross-sectional survey investigation. Journal of Affective Disorders, 274, 405–410.

32. Du, J., Dong, L., Wang, T., Yuan, C., Fu, R., Zhang, L., Liu, B., Zhang, M., Yin, Y., Qin, J., Bouey, J., Zhao, M., & Li, X. (2020). Psychological symptoms among frontline healthcare workers during COVID-19 outbreak in Wuhan. General Hospital Psychiatry, 67, 144–145.

33. Zhu, Z., Xu, S., Wang, H., Liu, Z., Wu, J., Li, G., Miao, J., Zhang, C., Yang, Y., Sun, W., Zhu, S., Fan, Y., Chen, Y., Hu, J., Liu, J., & Wang, W. (2020). COVID-19 in Wuhan: Sociodemographic characteristics and hospital support measures associated with the immediate psychological impact on healthcare workers. EClinicalMedicine, 24(100443), 100443.

34. Li, G., Miao, J., Wang, H., Xu, S., Sun, W., Fan, Y., Zhang, C., Zhu, S., Zhu, Z., & Wang, W. (2020). Psychological impact on women health workers involved in COVID-19 outbreak in Wuhan: a cross-sectional study. Journal of Neurology, Neurosurgery, and Psychiatry, 91(8), 895–897.

35. Dong, Z.-Q., Ma, J., Hao, Y.-N., Shen, X.-L., Liu, F., Gao, Y., & Zhang, L. (2020). The social psychological impact of the COVID-19 pandemic on medical staff in China: A cross-sectional study. European Psychiatry: The Journal of the Association of European Psychiatrists, 63(1), e65.

36. Zhang, W.-R., Wang, K., Yin, L., Zhao, W.-F., Xue, Q., Peng, M., Min, B.-Q., Tian, Q., Leng, H.-X., Du, J.-L., Chang, H., Yang, Y., Li, W., Shangguan, F.-F., Yan, T.-Y., Dong, H.-Q., Han, Y., Wang, Y.-P., Cosci, F., & Wang, H.-X. (2020). Mental health and psychosocial problems of medical health workers during the COVID-19 epidemic in China. Psychotherapy and Psychosomatics, 89(4), 242–250.

37. Zhang, S. X., Liu, J., Afshar Jahanshahi, A., Nawaser, K., Yousefi, A., Li, J., & Sun, S. (2020). At the height of the storm: Healthcare staff’s health conditions and job satisfaction and their associated predictors during the epidemic peak of COVID-19. Brain, Behavior, and Immunity, 87, 144–146.

38. Corbett, G. A., Milne, S. J., Mohan, S., Reagu, S., Farrell, T., Lindow, S. W., Hehir, M. P., & O’Connell, M. P. (2020). Anxiety and depression scores in maternity healthcare workers during the Covid-19 pandemic. International Journal of Gynaecology and Obstetrics: The Official Organ of the International Federation of Gynaecology and Obstetrics, 151(2), 297–298.

39. Rossi, R., Socci, V., Pacitti, F., Di Lorenzo, G., Di Marco, A., Siracusano, A., & Rossi, A. (2020). Mental health outcomes among frontline and second-line health care workers during the Coronavirus disease 2019 (COVID-19) pandemic in Italy. JAMA Network Open, 3(5), e2010185.

40. Di Tella, M., Romeo, A., Benfante, A., & Castelli, L. (2020). Mental health of healthcare workers during the COVID-19 pandemic in Italy. Journal of Evaluation in Clinical Practice, 26(6), 1583–1587.

41. Maciaszek, J., Ciulkowicz, M., Misiak, B., Szczesniak, D., Luc, D., Wieczorek, T., Fila-Witecka, K., Gawlowski, P., & Rymaszewska, J. (2020). Mental health of medical and non-medical professionals during the peak of the COVID-19 pandemic: A cross-sectional nationwide study. Journal of Clinical Medicine, 9(8), 2527.

42. Luceño-Moreno, L., Talavera-Velasco, B., García-Albuerne, Y., & Martín-García, J. (2020). Symptoms of posttraumatic stress, anxiety, depression, levels of resilience and burnout in Spanish health personnel during the COVID-19 pandemic. International Journal of Environmental Research and Public Health, 17(15), 5514.

43. Yildirim, T. T., Atas, O., Asafov, A., Yildirim, K., & Balibey, H. (2020). Psychological status of healthcare workers during the covid-19 pandemic. Journal of the College of Physicians and Surgeons--Pakistan: JCPSP, 30(6), 26–31.

44. Civantos, A. M., Byrnes, Y., Chang, C., Prasad, A., Chorath, K., Poonia, S. K., Jenks, C. M., Bur, A. M., Thakkar, P., Graboyes, E. M., Seth, R., Trosman, S., Wong, A., Laitman, B. M., Harris, B. N., Shah, J., Stubbs, V., Choby, G., Long, Q., … Rajasekaran, K. (2020). Mental health among otolaryngology resident and attending physicians during the COVID- 19 pandemic: National study. Head & Neck, 42(7), 1597–1609.

45. Evanoff, B. A., Strickland, J. R., Dale, A. M., Hayibor, L., Page, E., Duncan, J. G., Kannampallil, T., & Gray, D. L. (2020). Work-related and personal factors associated with mental well-being during the COVID-19 response: Survey of health care and other workers. Journal of Medical Internet Research, 22(8), e21366.

46. Liu, C.-Y., Yang, Y.-Z., Zhang, X.-M., Xu, X., Dou, Q.-L., Zhang, W.-W., & Cheng, A. S. K. (2020). The prevalence and influencing factors in anxiety in medical workers fighting COVID-19 in China: a cross-sectional survey. Epidemiology and Infection, 148 (e98), e98.

47. Kang, L., Ma, S., Chen, M., Yang, J., Wang, Y., Li, R., Yao, L., Bai, H., Cai, Z., Xiang Yang, B., Hu, S., Zhang, K., Wang, G., Ma, C., & Liu, Z. (2020). Impact on mental health and perceptions of psychological care among medical and nursing staff in Wuhan during the 2019 novel coronavirus disease outbreak: A cross-sectional study. Brain, Behavior, and Immunity, 87, 11–17.

48. Magnavita, N., Tripepi, G., & Di Prinzio, R. R. (2020). Symptoms in health care workers during the COVID-19 epidemic. A cross-sectional survey. International Journal of Environmental Research and Public Health, 17(14), 5218.

49. Xing, J., Sun, N., Xu, J., Geng, S., & Li, Y. (2020). Study of the mental health status of medical personnel dealing with new coronavirus pneumonia. PloS One, 15(5), e0233145.

50. Lam, S. C., Arora, T., Grey, I., Suen, L. K. P., Huang, E. Y.-Z., Li, D., & Lam, K. B. H. (2020). Perceived risk and protection from infection and depressive symptoms among healthcare workers in mainland China and Hong Kong during COVID-19. Frontiers in Psychiatry, 11, 686.

51. Shahzad, F., Du, J., Khan, I., Fateh, A., Shahbaz, M., Abbas, A., & Wattoo, M. U. (2020). Perceived threat of COVID-19 contagion and frontline paramedics’ agonistic behaviour: Employing a stressor-strain-outcome perspective. International Journal of Environmental Research and Public Health, 17(14).

52. Wu, K., & Wei, X. (2020). Analysis of psychological and sleep status and exercise rehabilitation of front-line clinical staff in the fight against COVID-19 in China. Medical Science Monitor Basic Research, 26, e924085.

53. Man, M. A., Toma, C., Motoc, N. S., Necrelescu, O. L., Bondor, C. I., Chis, A. F., Lesan, A., Pop, C. M., Todea, D. A., Dantes, E., Puiu, R., & Rajnoveanu, R.-M. (2020). Disease perception and coping with emotional distress during COVID-19 pandemic: A survey among medical staff. International Journal of Environmental Research and Public Health, 17(13).

54. Liang, Y., Chen, M., Zheng, X., & Liu, J. (2020). Screening for Chinese medical staff mental health by SDS and SAS during the outbreak of COVID-19. Journal of Psychosomatic Research, 133(110102), 110102.

55. Li, X., Yu, H., Bian, G., Hu, Z., Liu, X., Zhou, Q., Yu, C., Wu, X., Yuan, T.-F., & Zhou, D. (2020). Prevalence, risk factors, and clinical correlates of insomnia in volunteer and at home medical staff during the COVID-19. Brain, Behavior, and Immunity, 87, 140–141.

56. Zhu, S., Wu, Y., Zhu, C.-Y., Hong, W.-C., Yu, Z.-X., Chen, Z.-K., Chen, Z.-L., Jiang, D.- G., & Wang, Y.-G. (2020). The immediate mental health impacts of the COVID-19 pandemic among people with or without quarantine managements. Brain, Behavior, and Immunity, 87, 56–58.

57. Sun, D., Yang, D., Li, Y., Zhou, J., Wang, W., Wang, Q., Lin, N., Cao, A., Wang, H., & Zhang, Q. (2020). Psychological impact of 2019 novel coronavirus (2019-nCoV) outbreak in health workers in China. Epidemiology and Infection, 148(e96), e96.

58. Do Duy, C., Nong, V. M., Ngo Van, A., Doan Thu, T., Do Thu, N., & Nguyen Quang, T. (2020). COVID-19-related stigma and its association with mental health of health-care workers after quarantine in Vietnam. Psychiatry and Clinical Neurosciences, 74(10), 566–568.

59. Kannampallil, T. G., Goss, C. W., Evanoff, B. A., Strickland, J. R., McAlister, R. P., & Duncan, J. (2020). Exposure to COVID-19 patients increases physician trainee stress and burnout. PloS One, 15(8), e0237301.

60. Chen, Y., Zhou, H., Zhou, Y., & Zhou, F. (2020). Prevalence of self-reported depression and anxiety among pediatric medical staff members during the COVID-19 outbreak in Guiyang, China. Psychiatry Research, 288(113005), 113005.

61. Cheng, F.-F., Zhan, S.-H., Xie, A.-W., Cai, S.-Z., Hui, L., Kong, X.-X., Tian, J.-M., & Yan, W.-H. (2020). Anxiety in Chinese pediatric medical staff during the outbreak of Coronavirus Disease 2019: a cross-sectional study. Translational Pediatrics, 9(3), 231–236.

62. Liu, Y., Wang, L., Chen, L., Zhang, X., Bao, L., & Shi, Y. (2020). Mental health status of paediatric medical workers in China during the COVID-19 outbreak. Frontiers in Psychiatry, 11, 702.

63. Mo, Y., Deng, L., Zhang, L., Lang, Q., Liao, C., Wang, N., Qin, M., & Huang, H. (2020). Work stress among Chinese nurses to support Wuhan in fighting against COVID-19 epidemic. Journal of Nursing Management, 28(5), 1002–1009.

64. Cai, W., Lian, B., Song, X., Hou, T., Deng, G., & Li, H. (2020). A cross-sectional study on mental health among health care workers during the outbreak of Corona Virus Disease 2019. Asian Journal of Psychiatry, 51(102111), 102111.

65. Wong, K. C., Han, X. A., Tay, K. S., Koh, S. B., & Howe, T. S. (2020). The psychological impact on an orthopaedic outpatient setting in the early phase of the COVID-19 pandemic: a cross-sectional study. Journal of Orthopaedic Surgery and Research, 15(1), 322.

66. Khanna, R. C., Honavar, S. G., Metla, A. L., Bhattacharya, A., & Maulik, P. K. (2020). Psychological impact of COVID-19 on ophthalmologists-in-training and practising ophthalmologists in India. Indian Journal of Ophthalmology, 68(6), 994–998.

67. Szepietowski, J. C., Krajewski, P., Biłynicki-Birula, R., Poznański, P., Krajewska, M., Rymaszewska, J., & Matusiak, Ł. (2020). Mental health status of health care workers during the COVID-19 outbreak in Poland: One region, two different settings. Dermatologic Therapy, e13855.

68. Chen, X., Zhang, S. X., Jahanshahi, A. A., Alvarez-Risco, A., Dai, H., Li, J., & Ibarra, V. G. (2020). Belief in a COVID-19 conspiracy theory as a predictor of mental health and well-being of health care workers in Ecuador: Cross-sectional survey study. JMIR Public Health and Surveillance, 6(3), e20737.

69. Yang, S., Kwak, S. G., Ko, E. J., & Chang, M. C. (2020). The mental health burden of the COVID-19 pandemic on physical therapists. International Journal of Environmental Research and Public Health, 17(10), 3723.

70. Temsah, M.-H., Al-Sohime, F., Alamro, N., Al-Eyadhy, A., Al-Hasan, K., Jamal, A., Al-Maglouth, I., Aljamaan, F., Al Amri, M., Barry, M., Al-Subaie, S., & Somily, A. M. (2020). The psychological impact of COVID-19 pandemic on health care workers in a MERS-CoV endemic country. Journal of Infection and Public Health, 13(6), 877–882.

71. Xiao, H., Zhang, Y., Kong, D., Li, S., & Yang, N. (2020). The effects of social support on sleep quality of medical staff treating patients with Coronavirus disease 2019 (COVID-19) in January and February 2020 in China. Medical Science Monitor: International Medical Journal of Experimental and Clinical Research, 26, e923549.

72. Daugherty, A. M., & Arble, E. P. (2020). Prevalence of mental health symptoms in residential healthcare workers in Michigan during the covid-19 pandemic. Psychiatry Research, 291(113266), 113266.

73. Tu, Z.-H., He, J.-W., & Zhou, N. (2020). Sleep quality and mood symptoms in conscripted frontline nurse in Wuhan, China during COVID-19 outbreak: A cross-sectional study: A cross-sectional study. Medicine, 99(26), e20769.

74. Wang, S., Xie, L., Xu, Y., Yu, S., Yao, B., & Xiang, D. (2020). Sleep disturbances among medical workers during the outbreak of COVID-2019. Occupational Medicine (Oxford, England), 70(5), 364–369.

75. Teo, W. Z. Y., Soo, Y. E., Yip, C., Lizhen, O., & Chun-Tsu, L. (2020). The psychological impact of COVID-19 on “hidden” frontline healthcare workers. The International Journal of Social Psychiatry, 20764020950772.

76. Hines, S. E., Chin, K. H., Levine, A. R., & Wickwire, E. M. (2020). Initiation of a survey of healthcare worker distress and moral injury at the onset of the COVID-19 surge. American Journal of Industrial Medicine, 63(9), 830–833.

77. Huang, Y., & Zhao, N. (2020). Generalized anxiety disorder, depressive symptoms and sleep quality during COVID-19 outbreak in China: a web-based cross-sectional survey. Psychiatry Research, 288(112954), 112954.

78. Wang, W., Song, W., Xia, Z., He, Y., Tang, L., Hou, J., & Lei, S. (2020). Sleep disturbance and psychological profiles of medical staff and non-medical staff during the early outbreak of COVID-19 in Hubei Province, China. Frontiers in Psychiatry, 11, 733.

79. Zhan, Y., Liu, Y., Liu, H., Li, M., Shen, Y., Gui, L., Zhang, J., Luo, Z., Tao, X., & Yu, J. (2020). Factors associated with insomnia among Chinese front-line nurses fighting against COVID-19 in Wuhan: A cross-sectional survey. Journal of Nursing Management, 28(7), 1525–1535.

80. Zhang, C., Yang, L., Liu, S., Ma, S., Wang, Y., Cai, Z., Du, H., Li, R., Kang, L., Su, M., Zhang, J., Liu, Z., & Zhang, B. (2020). Survey of insomnia and related social psychological factors among medical staff involved in the 2019 novel Coronavirus disease outbreak. Frontiers in Psychiatry, 11, 306.

81. Asaoka, H., Koido, Y., Kawashima, Y., Ikeda, M., Miyamoto, Y., & Nishi, D. (2020). Post-traumatic stress symptoms among medical rescue workers exposed to COVID-19 in Japan. Psychiatry and Clinical Neurosciences, 74(9), 503–505.

82. Barello, S., Palamenghi, L., & Graffigna, G. (2020). Burnout and somatic symptoms among frontline healthcare professionals at the peak of the Italian COVID-19 pandemic. Psychiatry Research, 290(113129), 113129.

83. Wu, Y., Wang, J., Luo, C., Hu, S., Lin, X., Anderson, A. E., Bruera, E., Yang, X., Wei, S., & Qian, Y. (2020). A comparison of burnout frequency among oncology physicians and nurses working on the frontline and usual wards during the COVID-19 epidemic in Wuhan, China. Journal of Pain and Symptom Management, 60(1), e60–e65.

84. Osama, M., Zaheer, F., Saeed, H., Anees, K., Jawed, Q., Syed, S. H., & Sheikh, B. A. (2020). Impact of COVID-19 on surgical residency programs in Pakistan; A residents’ perspective. Do programs need formal restructuring to adjust with the “new normal”? A cross-sectional survey study. International Journal of Surgery (London, England), 79, 252–256.

85. Li, Z., Ge, J., Yang, M., Feng, J., Qiao, M., Jiang, R., Bi, J., Zhan, G., Xu, X., Wang, L., Zhou, Q., Zhou, C., Pan, Y., Liu, S., Zhang, H., Yang, J., Zhu, B., Hu, Y., Hashimoto, K.,… Yang, C. (2020). Vicarious traumatization in the general public, members, and non-members of medical teams aiding in COVID-19 control. Brain, Behavior, and Immunity, 88, 916–919.

86. Aafjes-van Doorn, K., Békés, V., Prout, T. A., & Hoffman, L. (2020). Psychotherapists’ vicarious traumatization during the COVID-19 pandemic. Psychological Trauma: Theory, Research, Practice and Policy, 12(S1), S148–S150.

87. Xu, J., Xu, Q.-H., Wang, C.-M., & Wang, J. (2020). Psychological status of surgical staff during the COVID-19 outbreak. Psychiatry Research, 288(112955), 112955.

88. Chen, H., Sun, L., Du, Z., Zhao, L., & Wang, L. (2020). A cross-sectional study of mental health status and self-psychological adjustment in nurses who supported Wuhan for fighting against the COVID-19. Journal of Clinical Nursing, 29(21–22), 4161–4170.

89. Pedrozo-Pupo, J. C., Pedrozo-Cortés, M. J., & Campo-Arias, A. (2020). Perceived stress associated with COVID-19 epidemic in Colombia: an online survey. Cadernos de Saude Publica, 36(5), e00090520.

90. Varshney, M., Parel, J. T., Raizada, N., & Sarin, S. K. (2020). Initial psychological impact of COVID-19 and its correlates in Indian Community: An online (FEEL-COVID) survey. PloS One, 15(5), e0233874.

91. Kiani Dehkordi, M., Sakhi, S., Gholamzad, S., Azizpour, M., & Shahini, N. (2020). Online Balint groups in healthcare workers caring for the COVID-19 patients in Iran. Psychiatry Research, 290(113034), 113034.

92. Preti, E., Di Mattei, V., Perego, G., Ferrari, F., Mazzetti, M., Taranto, P., Di Pierro, R., Madeddu, F., & Calati, R. (2020). The psychological impact of epidemic and pandemic outbreaks on healthcare workers: Rapid review of the evidence. Current Psychiatry Reports, 22(8), 43.

93. Maunder, R. G., Lancee, W. J., Balderson, K. E., Bennett, J. P., Borgundvaag, B., Evans, S., Fernandes, C. M. B., Goldbloom, D. S., Gupta, M., Hunter, J. J., McGillis Hall, L., Nagle, L. M., Pain, C., Peczeniuk, S. S., Raymond, G., Read, N., Rourke, S. B., Steinberg, R. J., Stewart, T. E., … Wasylenki, D. A. (2006). Long-term psychological and occupational effects of providing hospital healthcare during SARS outbreak. Emerging Infectious Diseases, 12(12), 1924–1932.

94. McAlonan, G. M., Lee, A. M., Cheung, V., Cheung, C., Tsang, K. W. T., Sham, P. C., Chua, S. E., & Wong, J. G. W. S. (2007). Immediate and sustained psychological impact of an emerging infectious disease outbreak on health care workers. Canadian Journal of Psychiatry. Revue Canadienne de Psychiatrie, 52(4), 241–247.

95. Kisely, S., Warren, N., McMahon, L., Dalais, C., Henry, I., & Siskind, D. (2020). Occurrence, prevention, and management of the psychological effects of emerging virus outbreaks on healthcare workers: rapid review and meta-analysis. BMJ (Clinical Research Ed.), 369, m1642.

96. Chong, M.-Y., Wang, W.-C., Hsieh, W.-C., Lee, C.-Y., Chiu, N.-M., Yeh, W.-C., Huang, O.-L., Wen, J.-K., & Chen, C.-L. (2004). Psychological impact of severe acute respiratory syndrome on health workers in a tertiary hospital. The British Journal of Psychiatry: The Journal of Mental Science, 185, 127–133.

97. Liu, X., Kakade, M., Fuller, C. J., Fan, B., Fang, Y., Kong, J., Guan, Z., & Wu, P. (2012). Depression after exposure to stressful events: lessons learned from the severe acute respiratory syndrome epidemic. Comprehensive Psychiatry, 53(1), 15–23.

98. Wu, P., Fang, Y., Guan, Z., Fan, B., Kong, J., Yao, Z., Liu, X., Fuller, C. J., Susser, E., Lu, J., & Hoven, C. W. (2009). The psychological impact of the SARS epidemic on hospital employees in China: exposure, risk perception, and altruistic acceptance of risk. Canadian Journal of Psychiatry. Revue Canadienne de Psychiatrie, 54(5), 302–311.

99. Borges, L. M., Barnes, S. M., Farnsworth, J. K., Bahraini, N. H., & Brenner, L. A. (2020). A commentary on moral injury among health care providers during the COVID-19 pandemic. Psychological Trauma: Theory, Research, Practice and Policy, 12(S1), S138–S140.

100. Williamson, V., Murphy, D., & Greenberg, N. (2020). COVID-19 and experiences of moral injury in front-line key workers. Occupational Medicine (Oxford, England), 70(5), 317–319.

101. Burchill, C. N. (2019). Critical incident stress debriefing: Helpful, harmful, or neither? Journal of Emergency Nursing: JEN: Official Publication of the Emergency Department Nurses Association, 45(6), 611–612.

102. Benedek, D. M., Fullerton, C., & Ursano, R. J. (2007). First responders: mental health consequences of natural and human-made disasters for public health and public safety workers. Annual Review of Public Health, 28(1), 55–68.

103. Terhakopian, A., & Benedek, D. M. (2007). Hospital disaster preparedness: mental and behavioral health interventions for infectious disease outbreaks and bioterrorism incidents. American Journal of Disaster Medicine, 2(1), 43–50.

104. Dieltjens, T., Moonens, I., Van Praet, K., De Buck, E., & Vandekerckhove, P. (2014). A systematic literature search on psychological first aid: lack of evidence to develop guidelines. PloS One, 9(12), e114714.

105. Benhamou, K., & Piedra, A. (2020). CBT-informed interventions for essential workers during the COVID-19 pandemic. Journal of Contemporary Psychotherapy, 1–9.

106. Ping, N. P. T., Shoesmith, W. D., James, S., Nor Hadi, N. M., Yau, E. K. B., & Lin, L. J. (2020). Ultra brief psychological interventions for COVID-19 pandemic: Introduction of a locally-adapted brief intervention for mental health and psychosocial support service. The Malaysian Journal of Medical Sciences: MJMS, 27(2), 51–56.

107. Francis, B., Juares Rizal, A., Ahmad Sabki, Z., & Sulaiman, A. H. (2020). Remote Psychological First Aid (rPFA) in the time of Covid-19: A preliminary report of the Malaysian experience. Asian Journal of Psychiatry, 54(102240), 102240.

108. Nash, W. P. (2011). US Marine Corps and Navy combat and operational stress continuum model: A tool for leaders. Combat and operational behavioral health, 107–119.

109. Albott, C. S., Wozniak, J. R., McGlinch, B. P., Wall, M. H., Gold, B. S., & Vinogradov, S. (2020). Battle Buddies: Rapid deployment of a Psychological Resilience Intervention for health care workers during the COVID-19 pandemic. Anesthesia and Analgesia, 131(1), 43–54.

110. Heath, C., Sommerfield, A., & von Ungern-Sternberg, B. S. (2020). Resilience strategies to manage psychological distress among healthcare workers during the COVID-19 pandemic: a narrative review. Anaesthesia, 75(10), 1364–1371.

111. Maunder, R. G., Leszcz, M., Savage, D., Adam, M. A., Peladeau, N., Romano, D., Rose, M., & Schulman, R. B. (2008). Applying the lessons of SARS to pandemic influenza: An evidence-based approach to mitigating the stress experienced by healthcare workers. Canadian Journal of Public Health. Revue Canadienne de Sante Publique, 99(6), 486–488.

112. Walton, M., Murray, E., & Christian, M. D. (2020). Mental health care for medical staff and affiliated healthcare workers during the COVID-19 pandemic. European Heart Journal. Acute Cardiovascular Care, 9(3), 241–247.

113. Wu, A. W., Connors, C., & Everly, G. S., Jr. (2020). COVID-19: Peer support and crisis communication strategies to promote institutional resilience. Annals of Internal Medicine, 172(12), 822–823.

114. Gentry, J. E., Baranowsky, A. B., & Dunning, K. (2002). The Accelerated Recovery Program (ARP) for compassion fatigue. In C. R. Figley (Ed.), Psychosocial stress series, no. 24. Treating compassion fatigue (p. 123–137). Brunner-Routledge.

115. Potter, P., Deshields, T., Berger, J. A., Clarke, M., Olsen, S., & Chen, L. (2013). Evaluation of a compassion fatigue resiliency program for oncology nurses. Oncology Nursing Forum, 40(2), 180–187.

116. Rajeswari, H., Sreelekha, B. K., Nappinai, S., Subrahmanyam, U., & Rajeswari, V. (2020). Impact of Accelerated Recovery Program on Compassion Fatigue among nurses in south India. Iranian Journal of Nursing and Midwifery Research, 25(3), 249–253.

117. Melnyk, B. M., Kelly, S. A., Stephens, J., Dhakal, K., McGovern, C., Tucker, S., Hoying, J., McRae, K., Ault, S., Spurlock, E., & Bird, S. B. (2020). Interventions to improve mental health, well-being, physical health, and lifestyle behaviors in physicians and nurses: A systematic review. American Journal of Health Promotion: AJHP, 34(8), 929–941.

118. Nochaiwong, S., Ruengorn, C., Awiphan, R., Ruanta, Y., Boonchieng, W., Nanta, S., Kowatcharakul, W., Pumpaisalchai, W., Kanjanarat, P., Mongkhon, P., Thavorn, K., Hutton, B., Wongpakaran, N., Wongpakaran, T., & Health Outcomes and Mental Health Care Evaluation Survey Research Group (HOME-Survey). (2020). Mental health circumstances among health care workers and general public under the pandemic situation of COVID-19 (HOME-COVID-19). Medicine, 99(26), e20751.

